# Assessing Factors Affecting Readiness for Reproductive, Maternal, Newborn, and Child Health Services, Ethiopia SPA Survey 2021-22

**DOI:** 10.64898/2026.03.11.26348195

**Authors:** Rachael Church, Fitsum Girma, Yunis Mussema, Gebeyehu Abelti, Abiy Shewarega

**Author notes:** Corresponding author:* Rachael Church.

## Abstract

This study assessed the readiness of health facilities across Ethiopia to provide essential reproductive, maternal, newborn, and child health (RMNCH) services. Using data from the 2021-22 Ethiopia Service Provision Assessment (ESPA), a comprehensive analysis of service readiness was conducted, employing principal component analysis (PCA) to identify the underlying factors affecting preparedness. Readiness scores were estimated for the critical RMNCH services of family planning, antenatal care (ANC), prevention of mother-to-child transmission of HIV (PMTCT), basic emergency obstetric and newborn care (BEmONC), comprehensive emergency obstetric care (CEmOC), child vaccination, and child curative care.

The analysis revealed significant variations in service readiness among regions and types of facilities. Referral hospitals, despite being at the top of the healthcare pyramid, often demonstrated lower readiness for certain services compared to lower-level facilities, suggesting potential challenges in resource allocation and prioritization. The PCA further highlighted the complex relationship between factors influencing service readiness, emphasizing the importance of resource availability (essential medicines, equipment, and supplies), presence of trained healthcare personnel, availability of WASH infrastructure and electricity, and availability of service guidelines.

These findings emphasize the need for a comprehensive approach to strengthen health systems in Ethiopia. We propose improving the availability of essential resources by investing in supply chain management, staff capacity building, and infrastructure (water and power access) development. Strategically investing in health system strengthening by addressing regional and other disparities is critical to achieving equitable access to quality RMNCH services.

## INTRODUCTION

### Background

The Ethiopia Service Provision Assessment (ESPA), conducted in 2021-22, is an instrumental tool for evaluating the readiness levels of facilities to provide Reproductive, Maternal, Newborn, and Child Health (RMNCH) services in Ethiopia over time.^1^

Globally, significant progress has been made in RMNCH outcomes over recent decades, resulting in notable reductions in maternal and child mortality rates. However, disparities remain, particularly in Sub-Saharan Africa. In 2020, countries in Sub-Saharan Africa accounted for 70% of global maternal deaths^2^ and over half of under-five child deaths,^3^ underscoring regional disparities in health outcomes Although these countries have generally expanded healthcare services and improved coverage of interventions like skilled birth attendance and vaccination, over the past 23 years, overall access to comprehensive RMNCH services remains limited.^3^ The Sustainable Development Goals (SDGs) set targets for child mortality for all countries to reduce the under-five mortality rate to 25 or fewer deaths per 1,000 live births by 2030, the neonatal mortality rate to 12 deaths or fewer per 1,000 live births by 2030 and the global maternal mortality ratio to less than 70 per 100,000 live births.^4^ Ethiopia’s successive Health Sector Development and Transformation Plans (HSTPs) and the recently launched Health Sector Development and Investment Plan, as well as the various Reproductive and Newborn Child health survival and development plans align with these global initiatives and aim to address these disparities by promoting universal health coverage and equitable access to RMNCH services.

### Reproductive, Maternal, Newborn, and Child Health (RMNCH) in Ethiopia

Ethiopia’s Demographic and Health Surveys (EDHS) show overall positive trends of RMNCH indicators over the past 20 years, although not at the rates required to meet the country’s HSTPs and the Sustainable Development Goals (SDGs). The U5MR has decreased from 123 deaths per 1000 births in 2005 to 51 deaths per 1000 births in 2024, but the SDG for U5MR is still half that value at 25 per 1000 births. ^5,6,7^ The neonatal mortality rate has decreased from 39 per 1000 births in 2005 to 25 per 1000 births in 2024, but the SDG is 12 per 1000 births.^5,6,7^ Modern contraceptive use among currently married women has increased from 14% in 2005 to 35% in 2024.^5,6^ However, the demand satisfied for family planning by modern methods was 67.7% in 2024, while the SDG for demand satisfied by modern methods is 75%.^6,7^ From 2005 to 2024, the maternal mortality ratio has declined from 673 per 100,000 live births to 141 per 100,000 live births - a great achievement, but still short of the SDG goal to reduce MMR to less than 70 maternal deaths per 100,000 live births.^5,6,7^ While these improvements in health outcomes are encouraging, there is still a long way to go to achieve the SDG targets.

### Service Readiness in Ethiopia

Health service readiness refers to the ability of health facilities to deliver essential health services effectively and consistently. Readiness indexes can encompass any number of critical factors, including the availability of trained health personnel, necessary medical equipment, essential medicines, diagnostics, and basic infrastructure such as running water, toilets, and electricity. In Ethiopia, the level of health service readiness is improving by several measures, but remains challenged by several factors. Interventions including healthcare worker training, supplying equipment, decentralization, and task sharing have shown promising improvements in the content, quality, and frequency of ANC visits in Ethiopia.^8,9^ However, infrastructure limitations, financial constraints, and workforce shortages are documented as remaining challenges for service readiness for RMNCH services.^10^ Health facilities, especially in rural areas, often lack essential medicines, medical supplies, and basic equipment, which undermines the quality of services. ^11^

A 2019 World Bank report highlighted that, by their readiness definition, only 51% of health centers in Ethiopia met the minimum requirements for service readiness in areas like maternal and child health, communicable diseases, and non-communicable disease management.^12^ Health workforce capacity is another critical issue, with a low ratio of healthcare providers to the population, often exacerbated by the uneven distribution of skilled professionals who prefer urban settings due to better living conditions and resources.^13^ Furthermore, political instability and limited budget allocations for health hinder sustainable improvements in readiness and response capabilities.^11^ Consequently, Ethiopia faces substantial challenges in achieving universal health coverage, necessitating targeted investments and policies to address these gaps effectively.

### Study Rationale

This study aims to contribute to our understanding of the readiness of health facilities to provide quality of reproductive, maternal, newborn and child health (RMNCH) primary care in Ethiopia. It uses a subset of variables informed by the six building blocks of health system strength identified by the World Health Organization (WHO), with a focus on service delivery.^13^

This study will help inform the health programs that work to improve the quality of RMNCH primary care in Ethiopia and policy makers to design and implement tailor made interventions addressing the gaps elaborated in the result. Improving service readiness is one of the key results of the activity/project. The assessment will support better implementation by pointing out the required capacity building support the health facilities need to improve service readiness. It will achieve this through the following objectives:

1. To estimate the readiness of health facilities to provide RMNCH services
2. To identify the factors that have the strongest effect on service readiness for RMNCH service providing facilities and make targeted recommendations for improving service readiness

## METHODS

### Data

This analysis uses data from the 2021-22 Ethiopia Service Provision Assessment (ESPA) survey. These surveys provide information about the readiness of health facilities to provide reproductive, maternal, newborn, child, and adolescent health (RMNCH) services, consisting of questions on health worker training and availability of service guidelines and the equipment and commodities necessary to provide the service. The ESPA includes eight different questionnaires: the facility inventory, the health provider interview, the antenatal care (ANC) client observation, the ANC client exit interview, the family planning (FP) client observation, the FP client exit interview, the sick child client observation, and the sick child caregiver exit interview.

Each survey followed a two-stage sampling design, with facilities selected from the national master facility list, and then health providers and clients selected from each selected facility. The facility sample included health centers, private clinics, and health posts and a census of hospitals in the country. This study used data from 1,158 facilities from the 2021-22 ESPA, of which 1044 provided family planning services, 544 provided ANC and PMTCT services, 650 provided normal delivery services, 341 provided Cesarean section services, 774 provided child vaccination services, and 1078 provided sick child care services.

We used the tracer items list from the WHO’s handbook on monitoring health systems^14^ to determine the list of services to consider under the RMNCH umbrella and generate a list of items needed for a facility to be considered “ready” to provide each of those services. The services selected from this handbook for this study based on their relevance to RMNCH needs include family planning, antenatal care (ANC), basic emergency obstetric and newborn care (BEmONC), comprehensive emergency obstetric care (CEmOC), child vaccination, child curative and preventative care, and prevention of mother to child transmission (PMTCT) of HIV. We combined ANC and PMTCT services into one group.

The tracer items for each of the six services, listed in Table 2, includes trained staff, service guidelines, and equipment and commodities needed to provide each service, as outlined in the WHO handbook. ^14^ We constructed variables for staff trained in providing each service within the 24 months prior to the survey as well as staff who had ever received training for that service. Guidelines were coded as either reported available and observed or not available. If guidelines were reported available but not observed, they were coded as not available. Similarly, for equipment to be coded as available, it must have been reported as available, observed, and shown to be functioning. Medicines and other commodities were coded as available if they were reported as available and at least one of the commodities was valid (not expired).

**Table 1:**
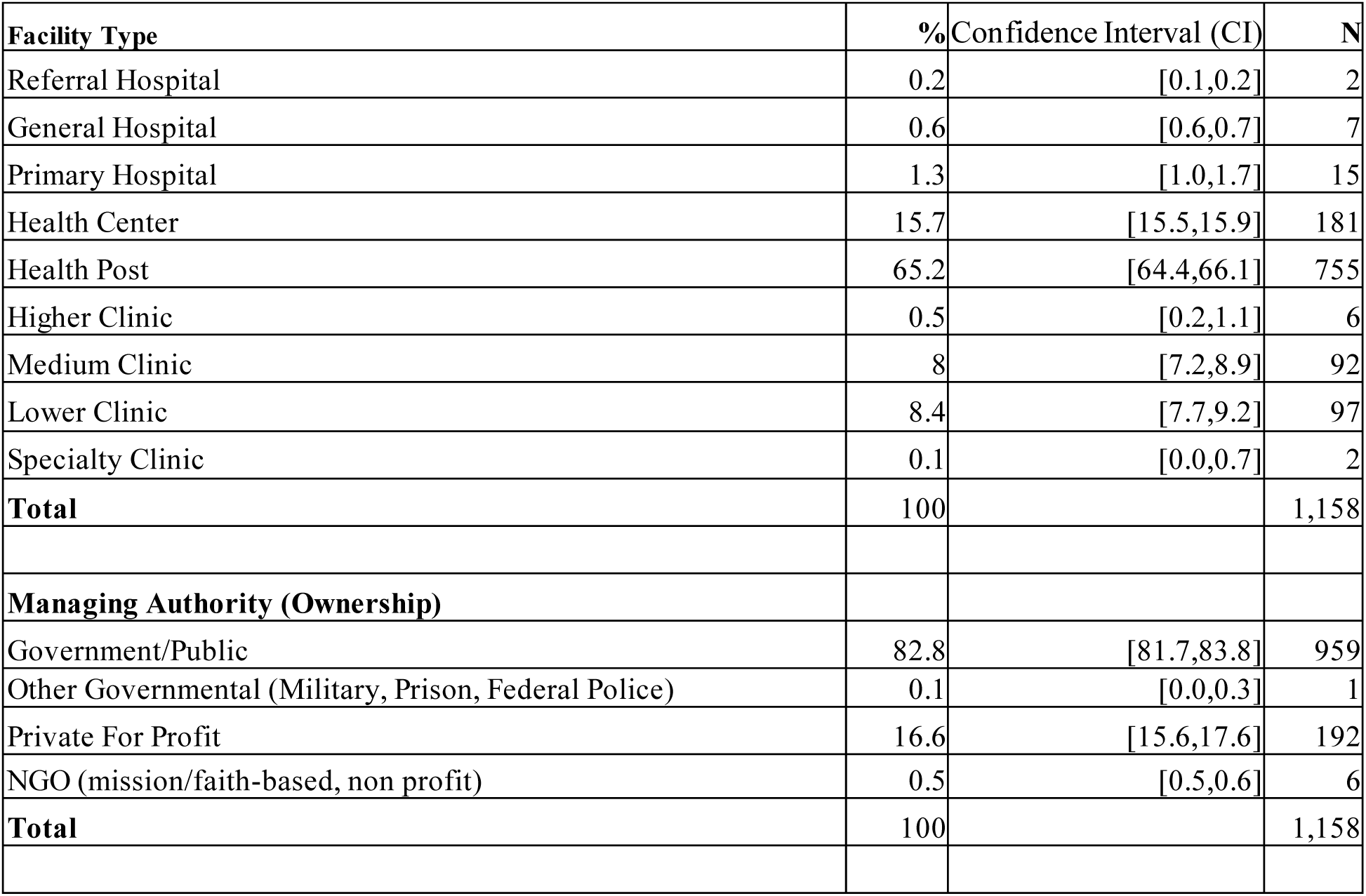
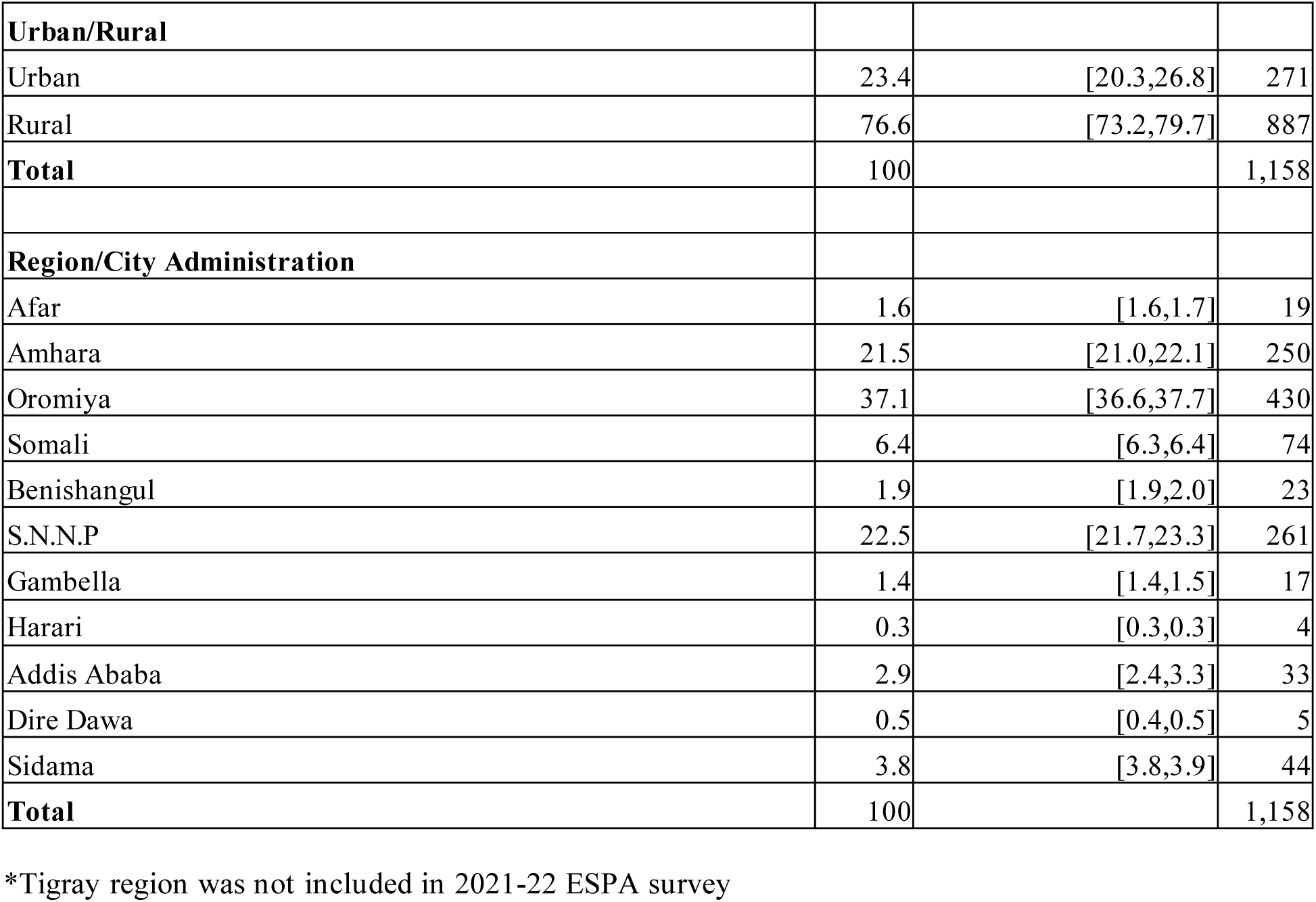
Distribution of facilities in Ethiopia by background characteristics (2021-22)

**Table 2:** Tracer Items for RMNCH Services

| Service | Tracer items |
| --- | --- |
| Family Planning | staff trained in family planning, family planning guidelines, combined oral contraceptive pills, injectable contraceptives, intrauterine contraceptive device (IUCD), implant, male condoms, blood pressure machine, stethoscope |
| ANC and PMTCT | staff trained in ANC, staff trained in providing PMTCT, staff trained in infant/young child feeding, ANC guidelines, PMTCT guidelines, infant/young child feeding guidelines, tetanus toxoid vaccine, blood pressure machine, stethoscope, hemoglobin tests, urine protein tests, urine glucose tests, syphilis tests, iron and folic acid tablets, visual and auditory privacy, dried blood spot testing, AZT, NVP, HIV tests |
| BEmONC | Staff trained in BEmONC, emergency transport, exam light, suction apparatus, manual vacuum extractor, vacuum aspirator/D&C kit, neonatal bag & mask, partograph, fetal stethoscope, newborn corner equipment, neonatal resuscitation kit, perinatal oxytocin, perinatal anticonvulsant, maternity waiting room gloves, antibiotic eye ointment, injectable uterotonic, injectable antibiotic, magnesium sulphate, intravenous solution with infusion set |
| CEmOC | staff trained in CEmOC, incubator, other external heat source, anesthesia machine, anesthesia machine w/ ventilator vaporisers & gas cylinders, anesthesia trolley, surgeon, anesthetist, blood transfusion available, blood typing capacity, blood obtained from national or regional blood bank OR blood obtained from other sources but screened for HIV and other transfusion transmissible infections, no shortage of blood in the last 3 months |
| Child vaccination | Staff trained in routine child immunization, guidelines on child immunization, refrigerator, cold box/vaccine carrier, sharps box, syringes and needles, vaccines |
| Child health services (curative and preventative) | staff trained in sick child curative care, staff trained in growth monitoring, guidelines on sick child care, guidelines on growth monitoring, oral rehydration salts, amoxicillin, cotrimoxazole, paracetamol, Vitamin A, mebendazole, zinc, child and infant scales, thermometer, stethoscope, growth charts, hemoglobin testing, malaria & stool microscopy, gentamycin, benzathine penicillin |

In most cases, variables were coded the same way for this analysis that they were for the ESPA 2021-22 final report. Examples of when this was not the case are detailed in the Analysis section.

### Analysis

First, we recorded all indicators included into binary variables. When preparing the dataset, we excluded five facilities that were sampled but declined to be interviewed, as the survey did not collect data on these facilities. We also excluded 144 facilities that were not functioning at the time of the survey. Generally, when coding variables, we followed the same methods as the ESPA final report, but there were some exceptions that caused our results to differ from the final report. These exceptions include:

- We did not exclude health posts where some final report tables did. If a facility claimed to provide a service, it was included in the analysis for that service regardless of the facility type.
- We did not exclude facilities that provided a service but did not regularly stock medication when creating the medication variables. All facilities that claimed to provide a service were included in the analysis for that service regardless of whether they regularly stocked medications.
- Similarly, for the child vaccination service, the final report only included facilities that reported regularly stocking vaccines in the denominator for vaccine variables, but we determined that a facility must routinely store vaccines to be ready to provide them, so all facilities that offered child vaccination were included, regardless of whether they regularly stocked vaccines.
- When coding variables for supplies and medications for a service, we did not count facilities that reported having those supplies/medications “available elsewhere in the facility” as having those supplies/medications because the validity of those supplies/medications was not assessed in the same survey question. Only medicines observed to be valid, and therefore in the service area, were included in this analysis.
- Some tables in the final report had equipment listed as available based on whether it was observed or reported, regardless of whether it was functional. Our analysis only coded an equipment variable as available if it was observed to be functioning.

We then used principal components analysis (PCA) to estimate the weight of each tracer item in predicting readiness for each service. PCA is a data reduction method that works by combining the fewest variables that account for the largest proportion of variance in, in this case, service readiness between facilities. Since PCA is designed to identify variance, it can interpret complex data without predefined weights or assumptions of linear relationships.^15^ We conducted a PCA for each of the six RMNCH services identified above and retained components with eigen values of at least 1.We considered a KMO value of 0.5 or higher to be valid.^16^

### Estimating Service Readiness

Though it is straightforward to identify reputable sources of indicators that must be included in an estimation of service readiness, there is no standard methodology for doing so. Mallick et.al. Compared several different methodologies for estimating quality of care for family planning services in 2017.^14^ In their analysis, they compared facility quality scores from simple additive, weighted additive, and PCA methodologies to determine the effects of the different approaches on the resulting service quality measures. In additive methodologies, indicators are coded into binary variables and grouped into domains with equal weight, creating a score based on how many points each indicator receives. PCA, on the other hand, is a variable reduction methodology, which combines sets of related variables into components and orders those components based on the amount of variance they each account for in the data. Mallick et al found that though PCA has more difficult indices to build and more challenging results to interpret than simple and weighted additive methods, the data reduction technique is still useful for calculating summary indices in analyses that include many variables. It determines how much weight each variable should have based on how it contributes to overall variation rather than using equal weighting systems. ^14^ The resulting indices allow comparison between facilities on service quality and readiness.

We predicted service readiness scores for each service based on the factor weights from the PCA. The factor loadings for each service are reported in Appendix Tables 1 through 6. We then classified facilities into tertiles based on their level of readiness and conducted Chi Squared tests to assess the statistical significance of the relationship between the readiness scores and facility characteristics. We tabulated the score groups by background characteristics to understand what types of facilities are most or least prepared to provide each service.

## RESULTS

### Service Readiness Estimation

Very high proportions of general hospitals, primary hospitals, and health centers all had low readiness for family planning services, with between 74% (health centers, CI 67-80) and 87% (primary hospitals, CI 81-92) of facilities having low readiness (Figure 1, Appendix Table 7). Conversely, only 10% (CI 4-25) of lower clinics had low readiness for family planning. Referral hospitals and higher clinics each only had two facilities included in the analysis for family planning services.

**Figure 1:**
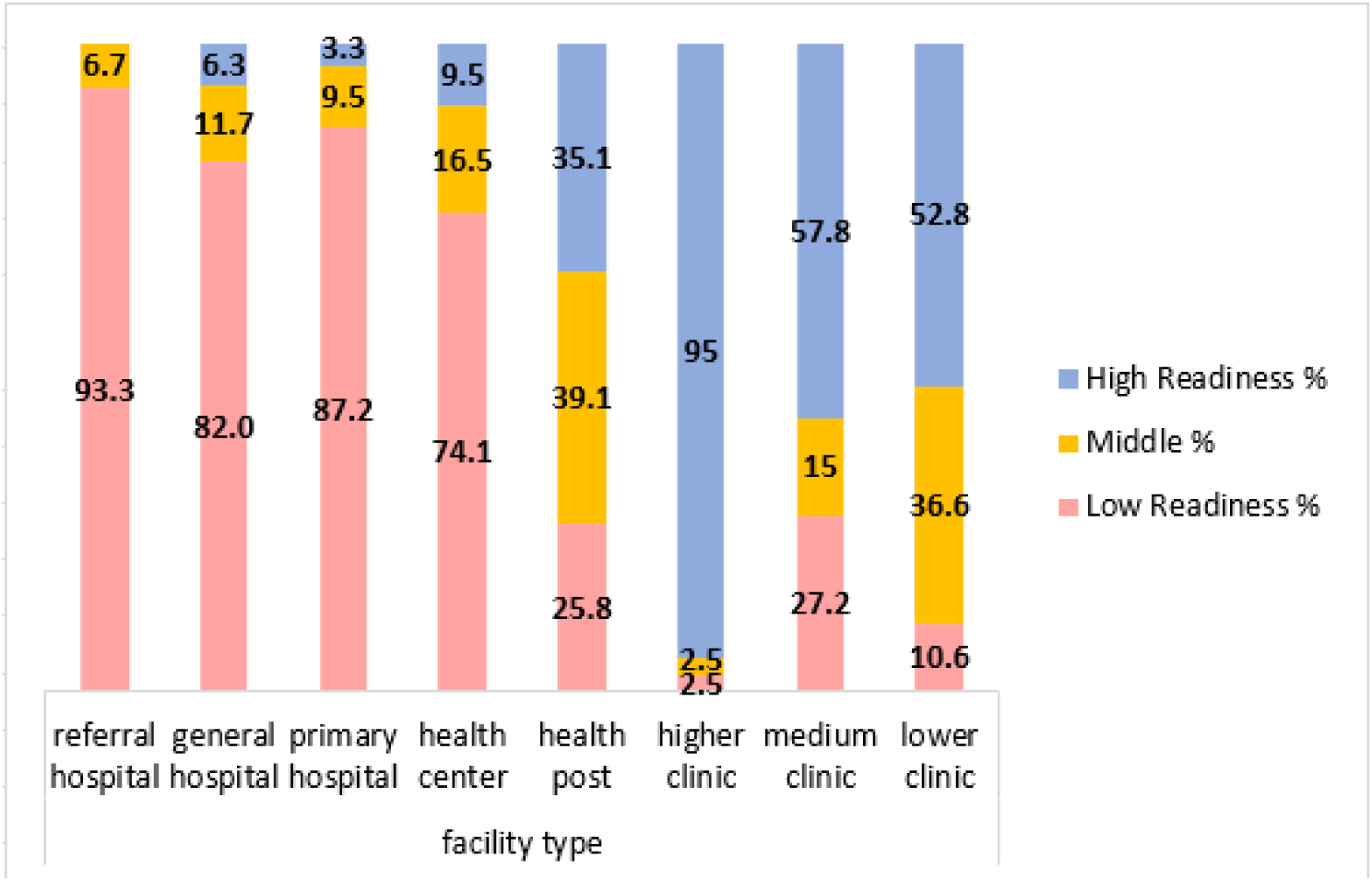
Service readiness estimates for family planning services by facility type in Ethiopia (2021-22)

The distribution of facility readiness by other facility characteristics can be seen in Appendix Table 7. Of note, privately managed facilities had the highest proportion of facilities with high readiness at 56% (CI 46-66) compared to other facility management types. Variation by urban/rural was limited for family planning facility readiness, though a higher proportion of urban facilities had low readiness (45%, CI 36-54) than did rural facilities (31%, CI 25-38). The distribution of facilities by region into score groups did not have enough eligible facilities in each region to produce statistically significant results. A total of 1,102 facilities were included in the family planning analysis.

Figure 2 and Appendix Table 8 show that only two regions had 50% or more of its facilities with a high readiness score: Sidama (60%, CI 45-73) and SNNP (50%, CI 33-68). There was strong variation between regions regarding service readiness for ANC and PMTCT, but Afar, Benishangul, Gambella, Harari, and Dire Dawa each only had 1-2 facilities that provided both ANC and PMTCT services, so results for these regions should be viewed with caution. After accounting for facility weights, only 166 out of 865 facilities reported offering both ANC and PMTCT. Despite these low numbers, the chi squared test for regions and facility readiness level was statistically significant (p=0.00), and are therefore presented and discussed here. The distribution of facility readiness by other facility characteristics can be seen in Annex Table 8. However, the chi-squared tests for the other facility characteristics were not statistically significant.

**Figure 2:**
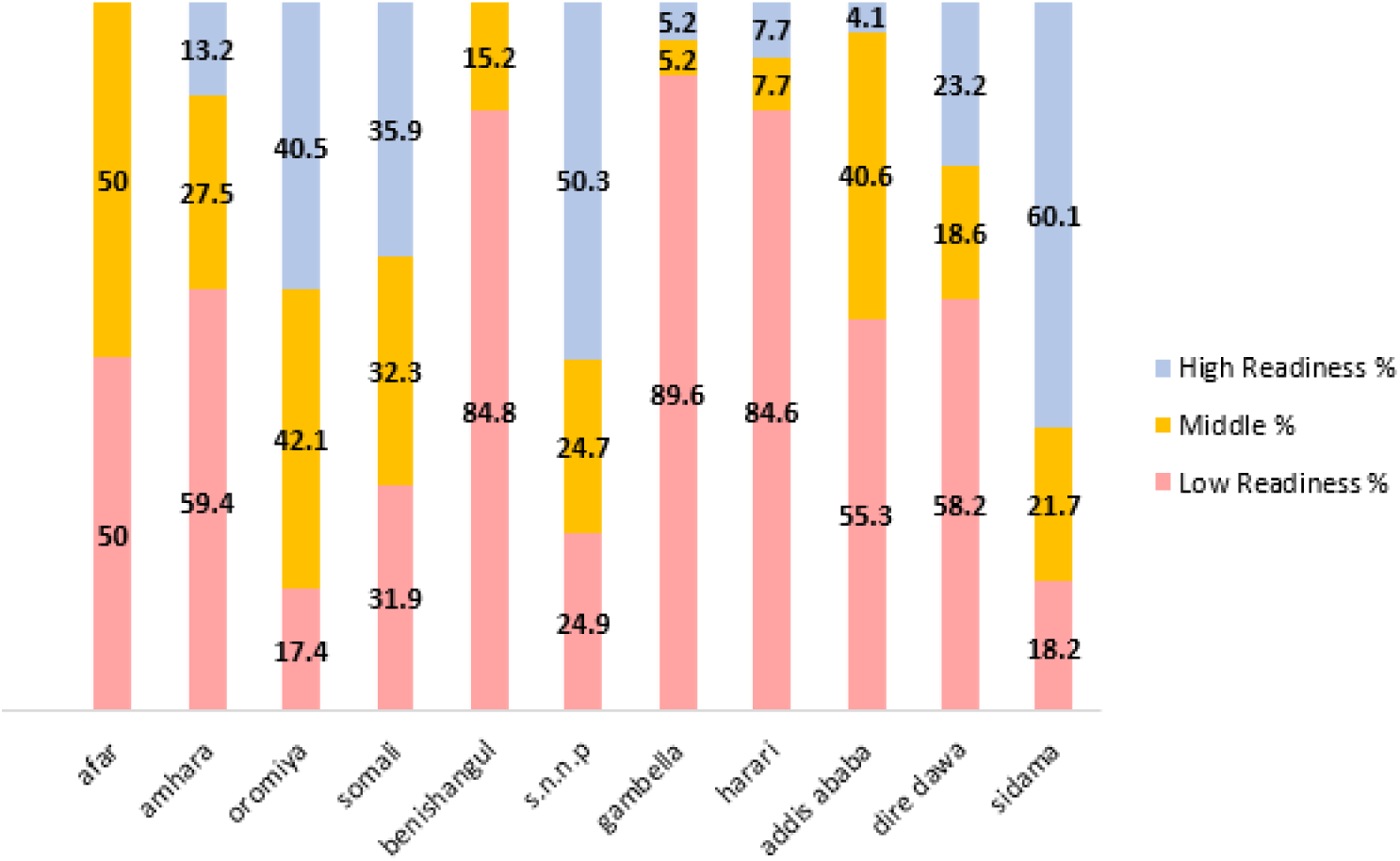
Service readiness estimates for ANC/PMTCT services by region in Ethiopia (2021-22)

In the BEmONC analysis, a total of 217 facilities were included. Appendix Table 9 shows that the facility types with the highest proportion of facilities scoring with a low readiness for BEmONC were general hospitals (84%, CI 77-90). In comparison, only 3.3% of medium sized clinics had a low readiness score for BEmONC (CI .9-11.6) (p=0.00). Additionally, 34% (CI 28-41) of publicly managed facilities had a low readiness score compared to 18% (CI 10-30) of privately managed facilities, (p=0.03). Figure 3 shows that 54% of Amhara facilities had low readiness (CI 40-68) compared to 13% of SNNP facilities (CI 7-23) (p=0.00). Afar, Benishangul, Gambella, Harari, and Dire Dawa regions all had fewer than 5 functioning facilities that offered BEmONC services.

**Figure 3:**
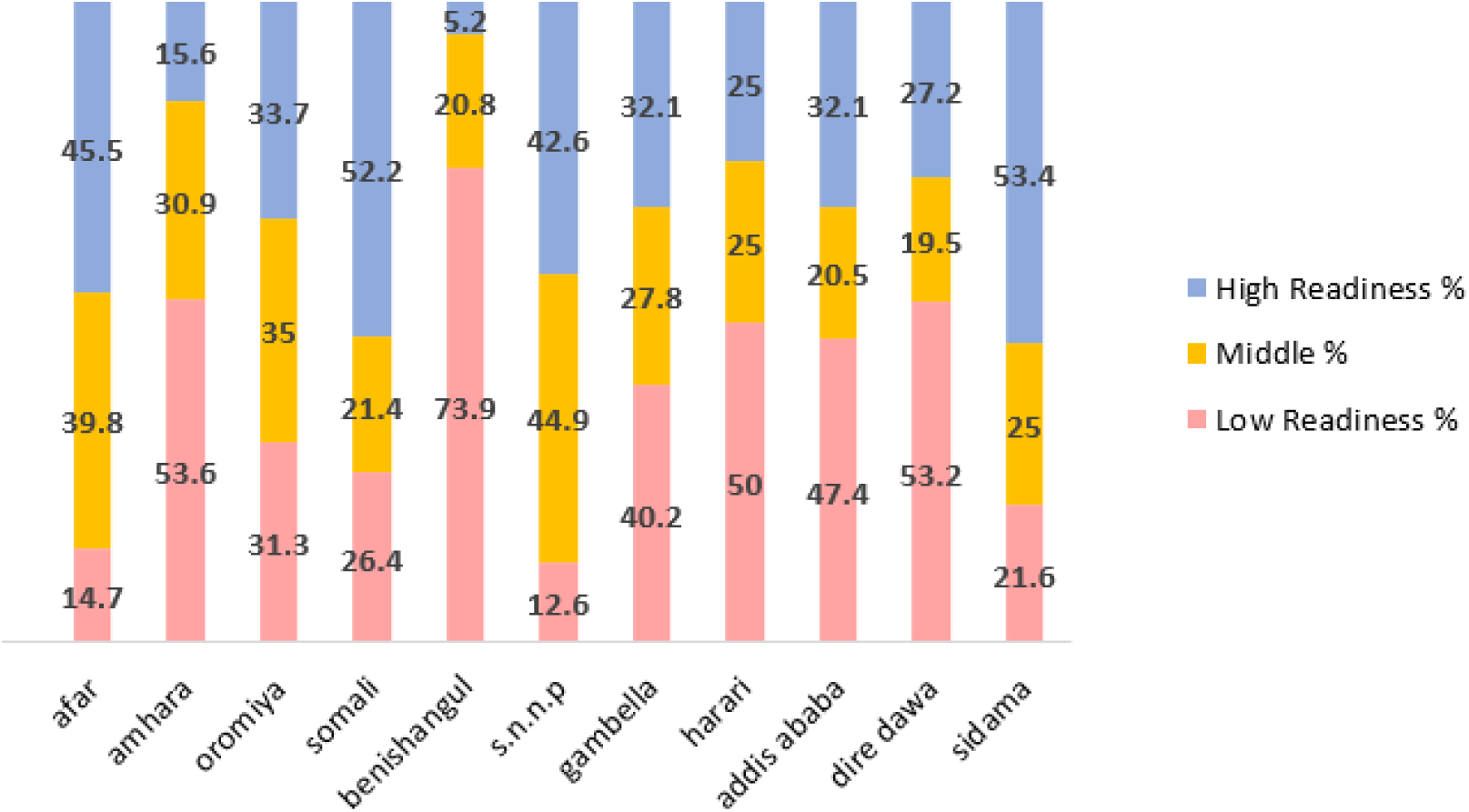
Service readiness estimates for BEmONC services in Ethiopia (2021-22)

After weighting, only 23 facilities offered C-section, so the sample size for this analysis was small (Appendix Table 10). A higher proportion of primary hospitals had higher readiness for CEmOC (41%, CI 29-55) compared to general hospitals (12%, CI 37-55) (p=0.005). Most private hospitals (66%, CI 39-86) had low readiness to provide CEmOC. While public facilities had a smaller proportion of facilities with low readiness (23%, CI 19-29), they still only had 39% (CI 33-44) of their facilities categorized as having high readiness (p=0.000). Figure 4 shows that urban facilities (39%, CI 27-54) have a higher proportion of facilities ranked as having low readiness compared to rural facilities (12%, CI 7-22), however only 4 total facilities that provide CEmOC services were located in rural areas (p=0.00).

**Figure 4:**
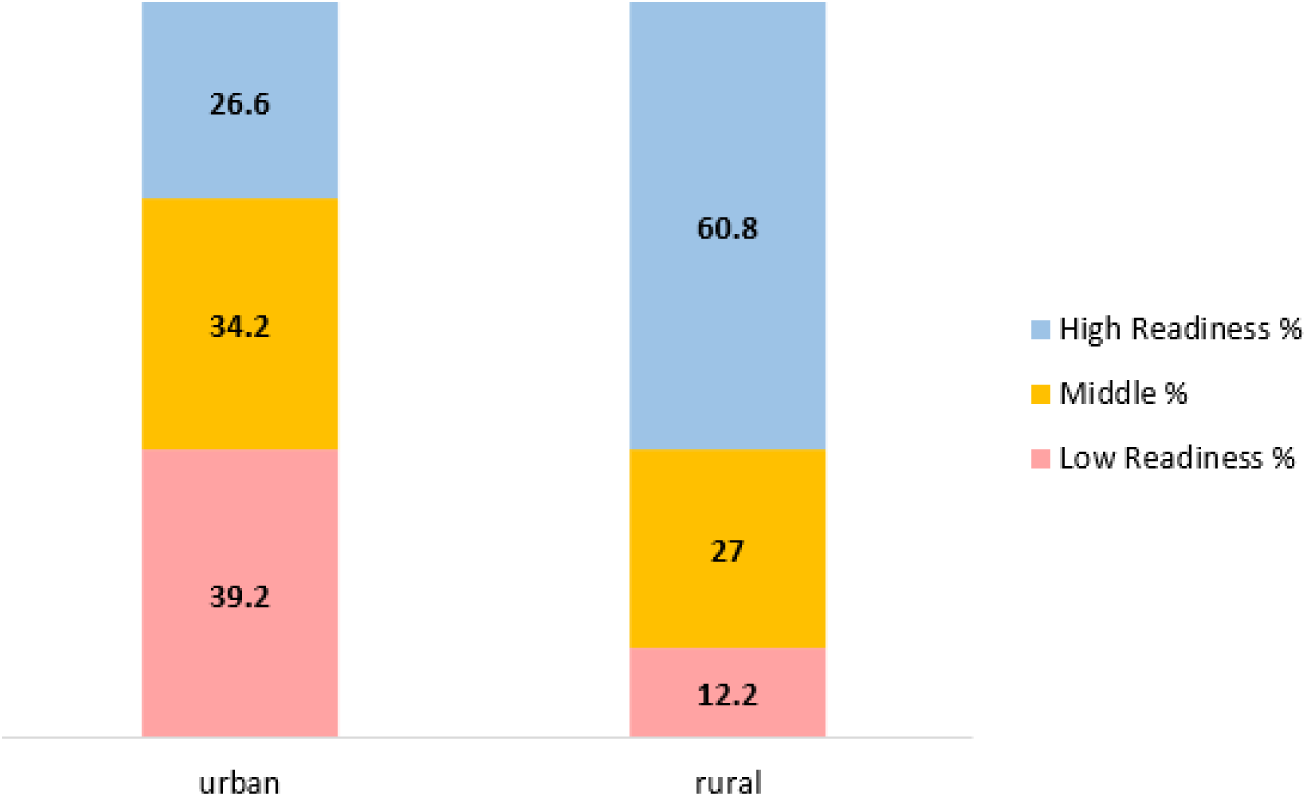
Service readiness estimates for CEmOC services in Ethiopia (2021-22)

A total of 891 facilities were included in the child vaccination analysis (Appendix Table 11). Among these, 702 were health posts and 172 were health centers. There were no medium, lower, or specialty clinics included in this analysis, and only one referral hospital. Health centers had the highest proportion of facilities categorized with low readiness for child vaccination (65%, CI 57-72), followed by general hospitals (57%, CI 47-67) and primary hospitals (52%, CI 45-60). Health posts had, by far, the highest proportion of facilities categorized as having high readiness, which was still only 33% (CI 26-42) (p=0.000). All facilities except two that were included in this analysis were public facilities. There was no statistically significant difference in the readiness levels between urban and rural facilities. Figure 5 shows that the region with the highest proportion of facilities categorized as having low readiness was Harari (68%, CI 49-82), but that this region had only two facilities included. Gambella (59%, CI 38-78), Benishangul (58%, CI 36-77), and Addis Ababa (54%, CI 36-71) regions all also had more than half of their facilities categorized as having low readiness. SNNP region had the lowest proportion of facilities in the low readiness category at 25% (CI 16-36) (p=0.019). 802 facilities were included in the analysis for child curative and growth monitoring services (Appendix Table 12). 96% of health centers had low readiness (CI 91-99). Comparatively, only 14% of health posts had low readiness (CI 10-20) (p=0.000). 795 of the facilities that provided this service were public facilities and only 24 were private. The difference between readiness levels of the different management authorities was not statistically significant. Figure 6 shows that 62% of urban facilities providing child curative care had low readiness (CI 47-75) compared to 28% of rural facilities (CI 23-33) (p=0.000).

**Figure 5:**
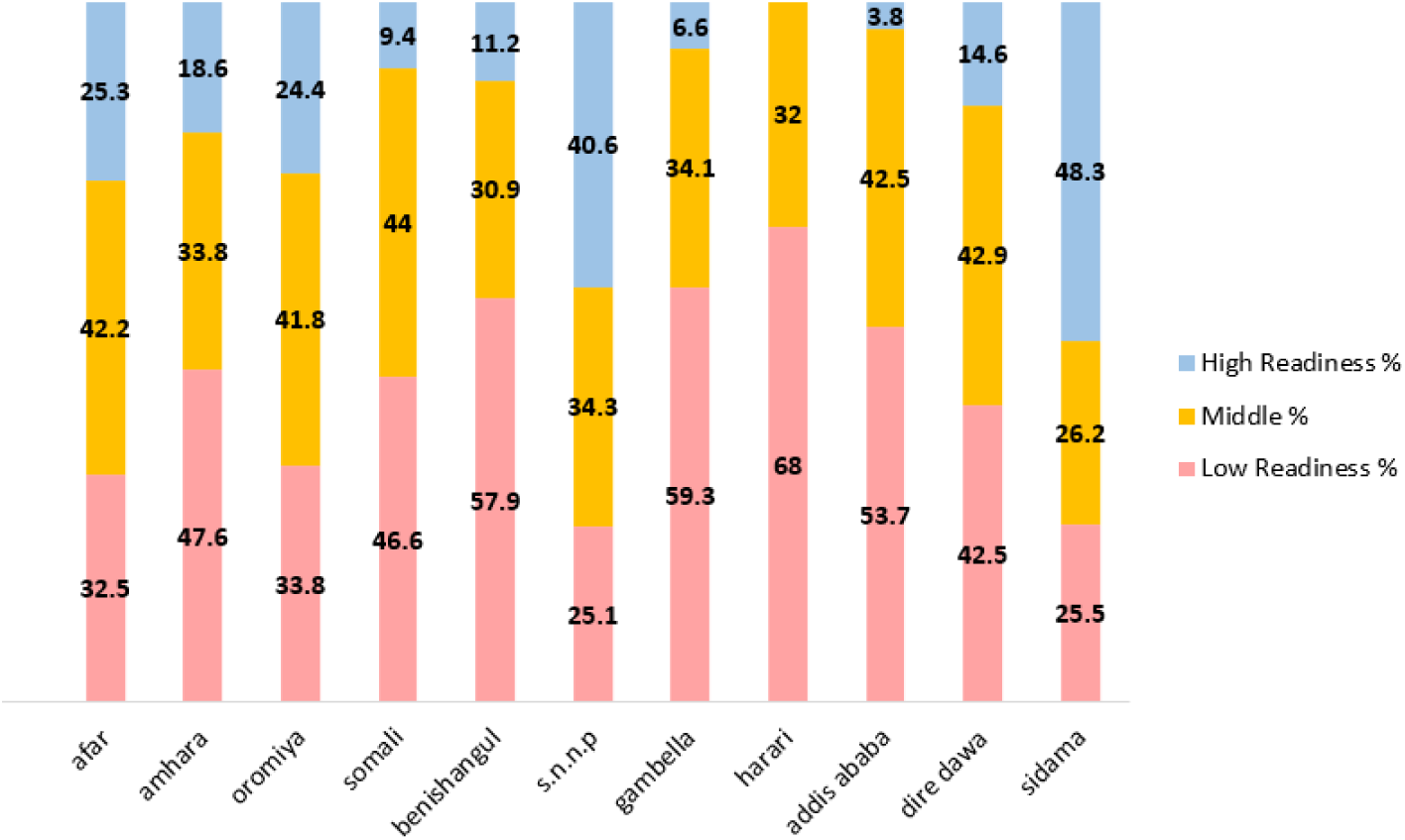
Service readiness estimates for child vaccination services in Ethiopia (2021-22)

**Figure 6:**
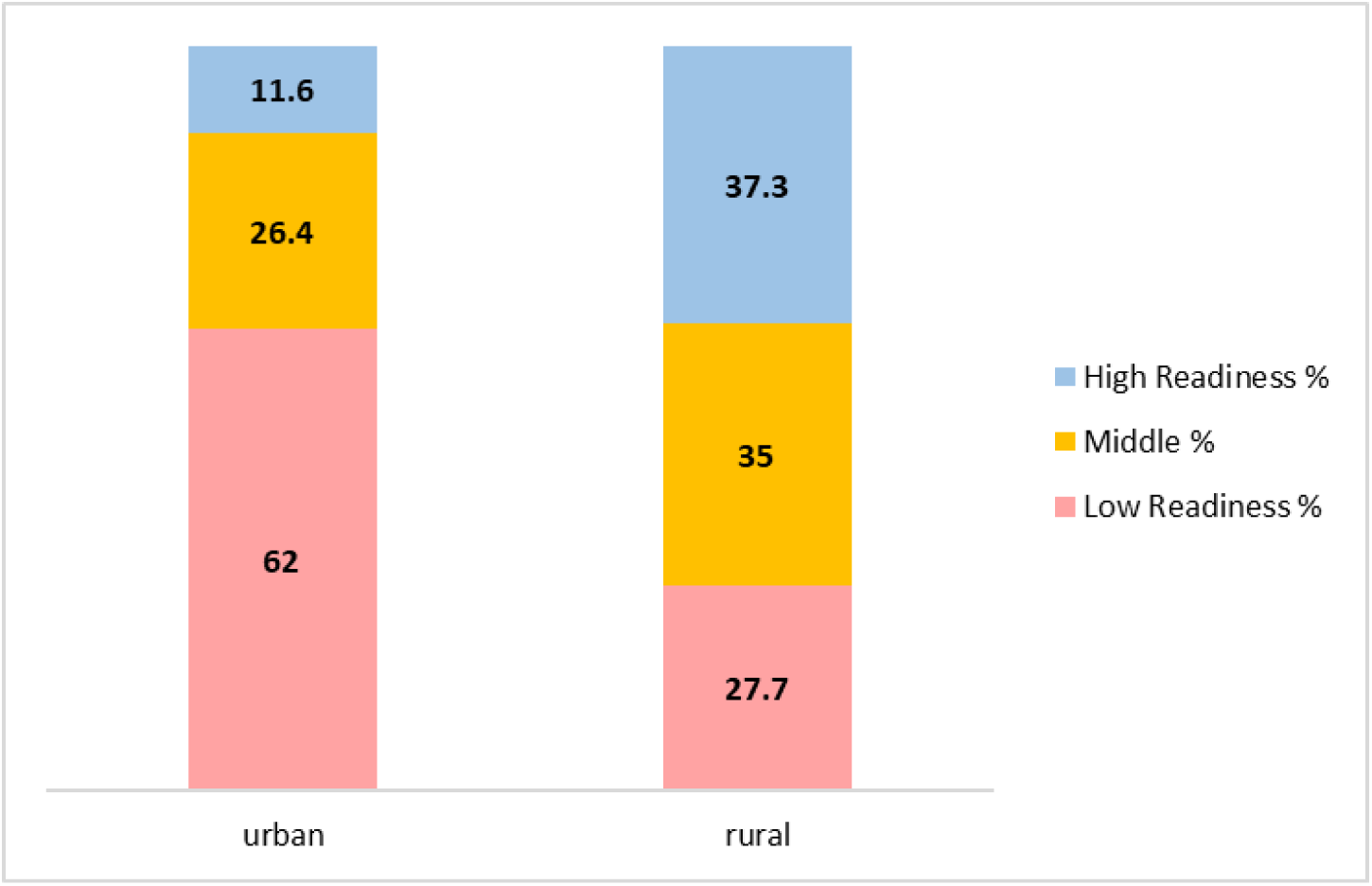
Service readiness estimates for child curative care and growth monitoring services in Ethiopia (2021-22)

Gambella, Harari, Addis Ababa, and Dire Dawa, each had fewer than 10 facilities that provided child curative care. One hundred percent (100%) of the Addis Ababa facilities included had low readiness. Afar (66%, CI 42-84), Somali (67%, CI 50-81), Harari (74%, CI 55-86), and Dire Dawa (63% CI 50-75) also had over half of their facilities categorized as having low readiness (p=0.002).

### Principal Component Analysis (PCA)

Tables 3-6 show the results from the PCAs, including the eigenvalues (the amount of variance in the data explained by each principal component), difference in eigenvalue between each principal component and the next, proportion of variance explained by the first three components, and the cumulative variance explained by the first three components for each service. The factor loadings for the first three components of each PCA can be seen in Appendix Tables 1-6. Each value in the table indicates the contribution (loading) of a specific variable to a particular component, with positive or negative values showing the direction of the relationship, and the magnitude indicating the strength. Accordingly, the availability of combined oral pill (0.464), injectable (0.468), implant (0.440), male condom (0.415) are dominant contributors for component1. This means that facilities with a higher presence of these items will have a score higher on this component. In component 2 availability of blood pressure monitors (fpbpm, 0.58) and stethoscopes (.fpsteth, 0.53) are dominant contributors, while staff trained in FP (0.55) is the major contributor in component 3.

**Table 3:** Principal component analysis output for service readiness indicators for family planning (2021-22)

| Component | Eigenvalue | Difference | Proportion | Cumulative |
| --- | --- | --- | --- | --- |
| Comp1 | 2.75393 | 0.940935 | 0.2504 | 0.2504 |
| Comp2 | 1.813 | 0.49778 | 0.1648 | 0.4152 |
| Comp3 | 1.31522 | 0.253055 | 0.1196 | 0.5347 |

**Table 4:**
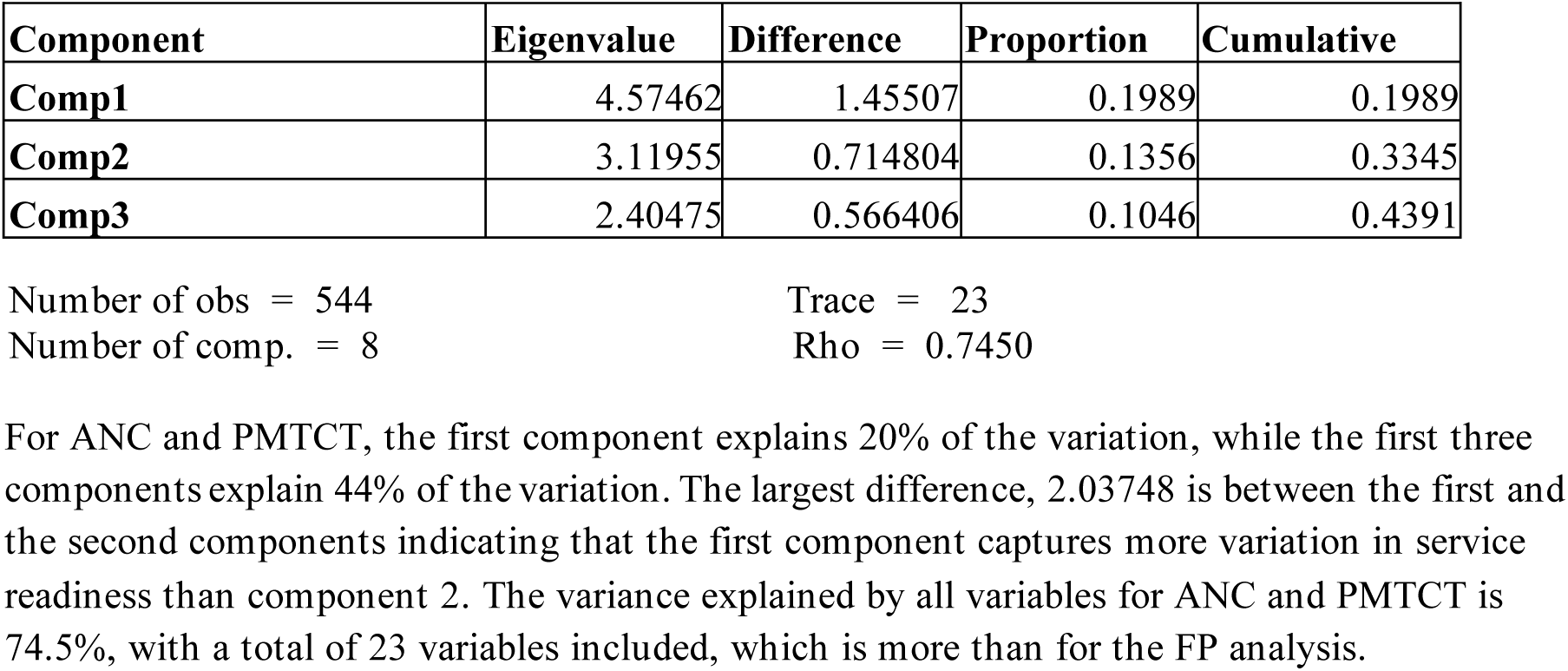
Principal component analysis output for service readiness indicators for ANC and PMTCT (2021-22)

**Table 5:**
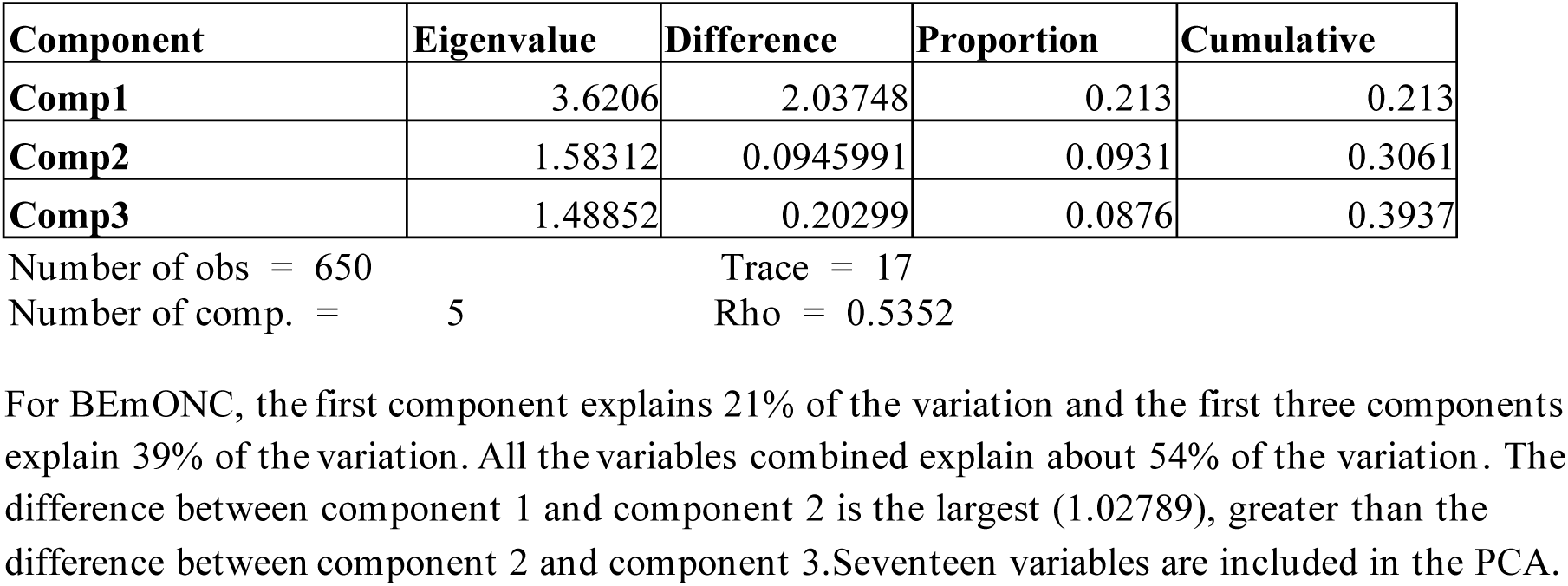
Principal component analysis output of BEmONC service readiness indicators (2021-22)

**Table 6:**
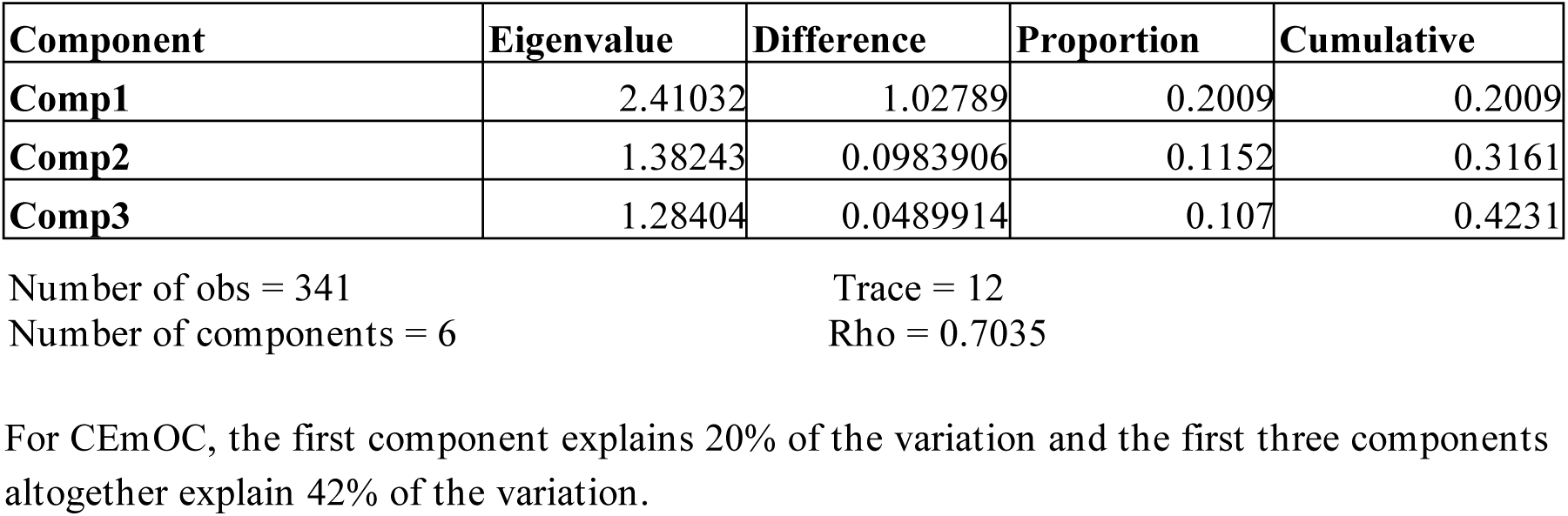
Principal component analysis output of CEmOC service readiness indicators (2021-22)

The eigenvalues in a PCA quantify how much of the data’s variance is captured by each principal component, helping to identify the most important features of the dataset whereas the difference indicates the reduction in eigenvalues between consecutive components. As shown in Table 3, the first component explains 25% of the variation, and the first three components together explain 53% of the variation in service readiness between facilities for family planning. The largest difference is between component1 and component2 (difference = 0.940935), showing that the first component captures more variance than the second. Rho, the proportion of total variance captured by all components is 63.13%. Trace explains the total number of variables included.

For ANC and PMTCT, the first component explains 20% of the variation, while the first three components explain 44% of the variation. The largest difference, 2.03748 is between the first and the second components indicating that the first component captures more variation in service readiness than component 2. The variance explained by all variables for ANC and PMTCT is 74.5%, with a total of 23 variables included, which is more than for the FP analysis.

For BEmONC, the first component explains 21% of the variation and the first three components explain 39% of the variation. All the variables combined explain about 54% of the variation . The difference between component 1 and component 2 is the largest (1.02789), greater than the difference between component 2 and component 3.Seventeen variables are included in the PCA.

For CEmOC, the first component explains 20% of the variation and the first three components altogether explain 42% of the variation.

For child vaccination, the first component explains 42% of the variation and the first three components explain 68% of the variation.

For child curative care, the first component explains 20% of the variation and the first three components explain 36% of the variation.

## DISCUSSION

The findings from the service readiness assessment for Ethiopia reveal several critical insights into the preparedness of health facilities to deliver essential maternal and child health services, particularly in the areas of family planning, ANC and PMTCT, BEmONC, CEmOC, child vaccination, and child curative care. These findings provide valuable information for policymakers, health system planners and program implementers to address the significant gaps in service delivery across different regions and facility types.

Principal Component Analysis (PCA) was employed to identify the underlying patterns and dimensions that contribute to service readiness across various health services in Ethiopia. By reducing the dimensionality of the data, PCA allowed us to distill complex factors into primary components that explain the majority of variance in service readiness. The PCA results reveal key factors influencing the preparedness of health facilities, providing more insight into the structural and functional elements that impact service delivery in Ethiopia’s health sector.

### Family Planning

The analysis (Appendix Table 7) indicates wide variation in readiness for family planning services by facility type, with about 75% or more of general hospitals, primary hospitals, and health centers having low readiness but less than 30% of health posts, medium clinics, and lower clinics having the same categorization.This stark contrast highlights an important challenge, as general, primary, and referral hospitals and health centers are expected to provide family planning services, yet they are clearly less equipped to deliver them. The Essential Health Services Package for Ethiopia^15^ puts family planning as a core component at the primary health care level, yet these providers of primary level care are seriously lacking in abilities. Privately managed facilities had a higher proportion of facilities with high readiness (56%) compared to public facilities (30%), a finding which is consistent with previous studies. For example, a study in Kenya found that private facilities often outperformed public ones in service delivery, particularly in family planning, due to better resource management and staffing levels. ^18^

For family planning services, the first principal component explained 25% of the variation in service readiness (Table 3). This dimension was primarily driven by the availability of contraceptive methods, which included combined oral pills, injectables, an d implants (Appendix Table 1). The concentration on contraceptive availability aligns with global health frameworks that emphasize the importance of supply-side factors, such as the availability of a wide range of contraceptive options, in ensuring high levels of family planning service readiness. These findings are consistent with similar studies in low- and middle-income countries (LMICs), where adequate supply of contraceptive methods remains a cornerstone for achieving service readiness.^19^

The fact that the availability of specific contraceptive methods, such as long-acting reversible contraception (LARC), played a central role in the PCA suggests that the demand for these methods, coupled with their effectiveness in preventing unwanted pregnancies, drives service delivery success. This resonates with findings from studies in Kenya^18^ and Nigeria,^20^ which identified similar supply-side constraints as limiting factors in the availability of family planning services.

### ANC and PMTCT

The readiness for ANC and PMTCT services (Appendix Table 8) showed a regional disparity, with regions such as Harari, Benishangul and Gambella exhibiting particularly low readiness (85%-90%) among only one or two facilities in each of these regions listed as providing these services to begin with. Sidama region had the highest proportion of facilities offering these services having high readiness at 60%. Facilities managed by private for profit show a larger proportion of facilities with high readiness for the ANC and PMTCT service delivery at 65% compared to government and publicly managed facilities, of which only 30% had high readiness for these services. These results suggest that regional and facility management factors play a significant role in service delivery, aligning with findings from the 2020 Ethiopia Health Sector Transformation Plan, which noted uneven resource distribution across regions.^12^ Similar trends have been observed in studies in other sub-Saharan African countries. In South Africa, rural and marginalized regions tended to have lower service readiness for ANC and PMTCT, for example.^21^ In Ethiopia, the disparity between regions is not so clear cut and consistent as being an urban or a rural issue, but some regions are demonstrably less resourced than others. The limited number of facilities offering both ANC and PMTCT services complicates comparisons and highlights an area for improvement: Afar, Benishangul, Gambella, Harari, and Dire Dawa regions all had two facilities or fewer that provided both ANC and PMTCT. Also, the fact that only 20% of the facilities assessed provide both ANC and PMTCT shows a lost opportunity to avail critical life saving integrated services.

For ANC and PMTCT services, the first principal component accounted for 20% of the variation in service readiness (Table 4). This dimension was heavily influenced by the availability of trained healthcare personnel, adherence to clinical guidelines for maternal and child health, and the availability of various tests (Appendix Table 2). These factors underline the critical role of human resources, standardized practices, and supplies in maintaining high -quality maternal care.

The strong association between service readiness and the presence of trained personnel is consistent with findings from other African countries, where the training and continuous professional development of health workers were identified as key contributors to the quality of maternal health services.^21^ Similarly, the emphasis on clinical guidelines in the PCA reflects global health recommendations, including those from the WHO, which have highlighted the importance of standardized protocols and evidence-based practices in reducing maternal and neonatal morbidity and mortality.^22^

Moreover, the importance of clinical guidelines in the PCA supports the findings of other studies, such as those conducted in Tanzania and Uganda, where health facilities with better adherence to clinical protocols saw higher service delivery success rates for ANC and PMTCT.^23, 24^ These studies also point out that adherence to guidelines is critical in resource-limited settings where healthcare personnel may have varying levels of experience and training.

### BEmONC and CEmOC

The results for BEmONC readiness (Appendix Table 9) revealed that service readiness varied by facility characteristics. A high proportion of general hospitals had low service readiness for BEmONC services (84%), whereas medium-sized clinics had a much lower proportion of facilities with low readiness (3%). This discrepancy likely stems from resource allocation and specialized training in higher-level facilities, which have more staff but also face higher patient loads and more logistical challenges. ^24^ Readiness also varied by the managing authority of the facility - public facilities had a higher proportion of low readiness for BEmONC (34.3%) compared to private facilities (18%), which echoes the trends observed in other settings, such as Tanzania, where public hospitals often struggle with resource constraints and management inefficiencies compared to private institutions.^25^

For CEmOC, primary hospitals had a higher proportion of facilities with high readiness (41%) than general hospitals (12%) (Appendix Table 10). This suggests that smaller facilities may be more nimble and able to maintain higher readiness levels with fewer resources. The observation that urban facilities had more than double the proportion of low readiness compared to rural facilities (39% vs. 12%) is consistent with the findings of a study in Uganda, which highlighted the challenge of urban congestion and overburdened infrastructure in metropolitan areas.^23^ This observation should be interpreted with caution, however, because there were only 4 rural facilities that provided CEmOC services.

The PCA for BEmONC and CEmOC services revealed that the first principal component accounted for a substantial proportion of the variation in service readiness (21% for BEmONC and 20% for CEmOC, Tables 5 and 6). For both services, the most influential factors were the presence of essential drugs and equipment for managing obstetric and neonatal emergencies.

This finding underscores the importance of equipping facilities with the necessary drugs and supplies to handle obstetric and neonatal emergencies, which are common causes of maternal and child mortality in LMICs. A study in Tanzania highlighted similar challenges, where facilities with inadequate resources and poorly equipped delivery rooms often struggle to provide essential emergency obstetric care, leading to poorer health outcomes.^25^ This is a critical area for improvement, as evidence from Kenya and Uganda shows that investing in equipment and improving supply chains can significantly enhance emergency care services.^18, 23^

### Child Vaccination and Child Curative Services

Health posts were found to have the highest proportion of facilities with high readiness to provide child vaccination services at 33% (Appendix Table 11), which is still relatively low. In comparison, fewer than 2% each of health centers, general hospitals, and primary hospitals had high service readiness for child vaccination. The facilities in Gambella and Benishangul regions both had more than 57% of facilities with low readiness, suggesting that factors such as regional funding allocation, training, and infrastructure development limit service delivery in these areas.

The results for child curative and growth monitoring services indicated a concerning 96% of health centers with low readiness (Appendix Table 12). This finding, consistent with studies in South Africa, indicates that health centers are often overwhelmed with high patient volumes and insufficient resources, resulting in suboptimal service delivery.^26^ The stark contrast with health posts, where only 14% of facilities had low readiness, suggests that smaller, more localized services may be more effective at addressing basic health needs, though they still face challenges in scaling up service delivery.

For child vaccination services, the first component of the PCA accounted for 42% of the variation in service readiness (Table 7), and the analysis identified infrastructure and the availability of vaccines as the primary drivers of service readiness. This indicates a persistent issue in equipping facilities to deliver routine immunization services effectively. This finding is consistent with similar studies in sub-Saharan Africa, where logistical challenges, such as the cold chain maintenance and vaccine storage, often hinder the successful delivery of immunization programs.^20^ In rural and remote areas, the lack of reliable electricity and transportation networks exacerbates these challenges, leading to lower vaccination coverage and readiness scores.

**Table 7:**
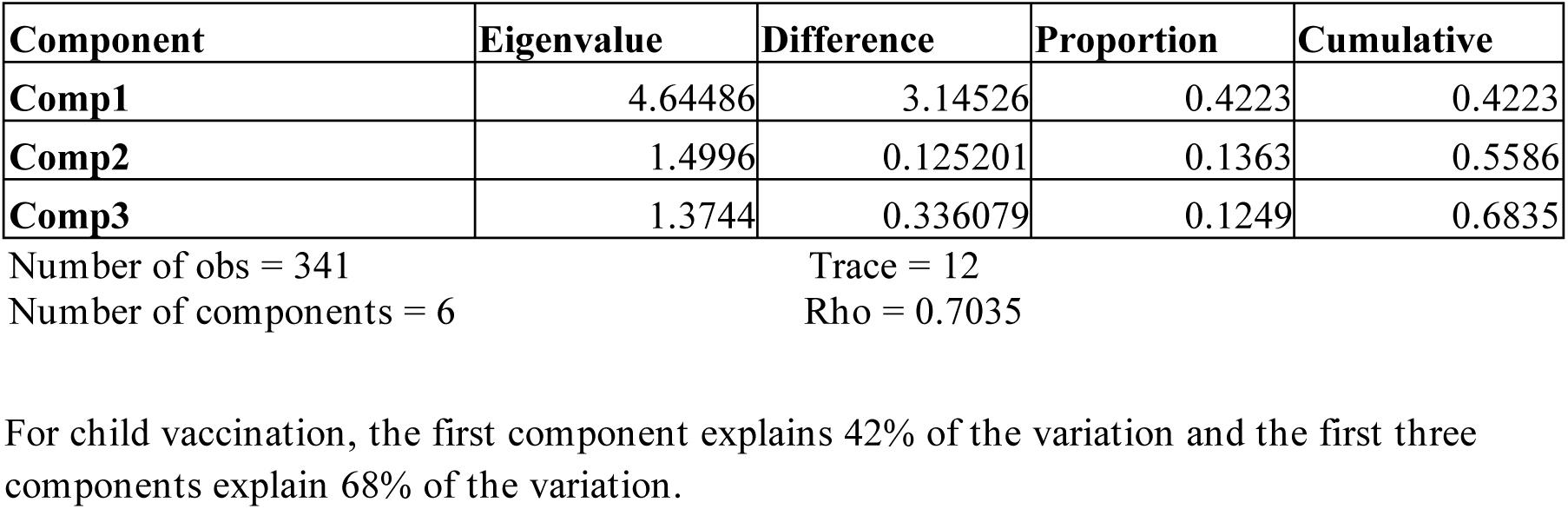
Principal component analysis output of child vaccination service readiness indicators (2021-22)

For child curative care services, the first component accounted for 20% of the variance (Table 8). Readiness for child curative services was most strongly driven by the availability of medications and testing equipment. Similar findings were observed in South Africa, where the lack of basic curative supplies limited the effectiveness of child health services in primary healthcare settings.^26^

**Table 8:**
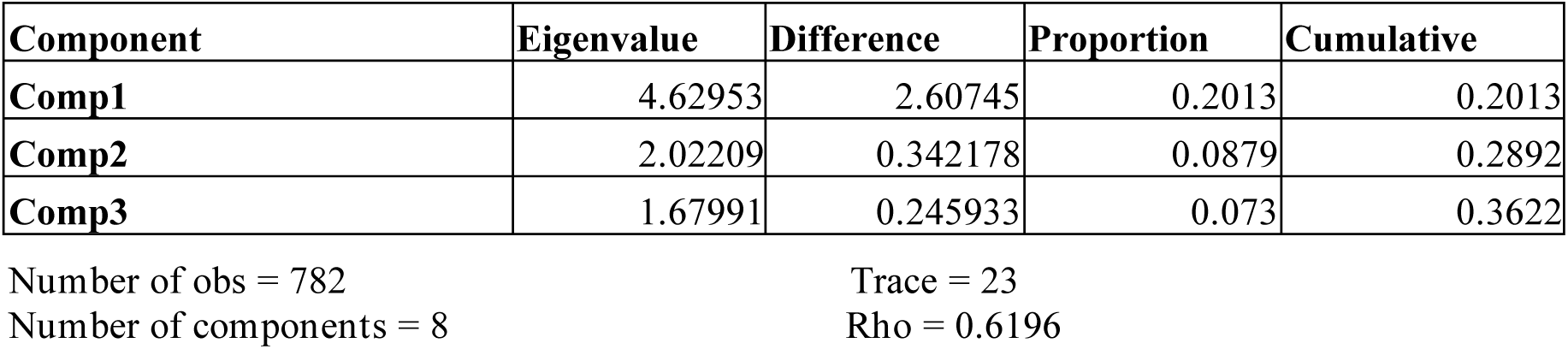
Principal component analysis output of child curative care service readiness indicators (2021-22)

### Implications of PCA Results

The PCA results provide critical insights into the primary drivers of service readiness in Ethiopia’s health facilities. The findings highlight the importance of addressing the supply-side factors—such as availability of drugs, equipment, and trained personnel—as well as strengthening adherence to clinical guidelines and protocols. These factors, which emerged as significant contributors to service readiness in the PCA, should be prioritized in future health system strengthening efforts.

Additionally, the regional variations uncovered in the PCA point to the need for targeted interventions based on local health system constraints. Regions with lower service readiness may require specific investments in training, infrastructure, and supply chain management to bridge the readiness gap. For example, regions such as Gambella and Harari, which were identified as having only one to two facilities each that could provide ANC, PMTCT, and BEmONC services, would especially benefit from region - specific resource mobilization and training initiatives.

## RECOMMENDATIONS

1. Strengthen supply chains for family planning and maternal health services: It is crucial to ensure the continuous availability of essential family planning commodities and maternal health supplies. The establishment of resilient and reliable supply chain systems will prevent stockouts and ensure consistent service delivery.
2. Enhance training and capacity building for healthcare providers: Continuous professional development and training for healthcare providers, especially in underserved and rural areas, should be prioritized to improve service delivery especially integration of services like ANC and PMTCT in all facilities and ensure adherence to clinical protocols.
3. Improve Infrastructure and Equipment for Emergency Obstetric and Neonatal Care: It is essential to upgrade health facility infrastructure and ensure the availability of necessary equipment for the provision of emergency obstetric and neonatal care services. This will improve outcomes in maternal and newborn health.
4. Ensure Adherence to Clinical Guidelines and Protocols: Efforts should be made to strengthen adherence to national clinical guidelines and protocols in maternal and child health services. This will improve the quality of care and reduce maternal and neonatal morbidity and mortality.
5. Targeted Resource Allocation for Underserved Regions: Allocate additional resources to regions that exhibit lower service readiness scores. This targeted investment will address disparities and improve healthcare access and quality in underserved areas.
6. Improve Data Collection and Monitoring Systems: Strengthening health information systems to enable real-time monitoring of service readiness and the effective management of health data will be vital in tracking progress and identifying gaps.

By addressing these recommendations, stakeholders can contribute to improving the service readiness and quality of maternal and child health services in Ethiopia, ultimately leading to better health outcomes across the country.

### Limitations

As mentioned in the methods section, the readiness scores calculated for this study are different than they would have been if all variables were coded the same way they were for the final report. These decisions were made to preserve consistency within the analyses and create estimates that were of the most relevance to policies and programs in Ethiopia. The reasons and details about these differences are noted in the methods section. Another analysis could repeat this study with the variables coded the same way as the final report. For example, another study could look into the difference in the readiness scores between facilities that have the necessary supplies and medications in the service area compared those reported elsewhere to explore the effect of this exclusion on the readiness scores For example, fewer facilities that provide ANC but not PMTCT have tetanus vaccines than facilities that offer both. By only looking at facilities that provide both ANC and PMTCT, we are overlooking a group of facilities that may be less ready to provide ANC than the group we’ve specified.

PCA does not allow us to directly compare readiness estimations between surveys because it will not use the variables to construct the components in the same way over multiple analyses.

The Tigray region of Ethiopia was not included in the 2021-22 SPA due to the northern conflict. This may render this research to be less generalizable for national performance in readiness.

## CONCLUSIONS

The findings from this study underscore the significant regional and facility-type disparities in service readiness across Ethiopia. Public health facilities, particularly in urban and underserved rural areas, face considerable challenges in meeting service readiness standards. This is compounded by issues in staffing, resource allocation, and infrastructure limitations. Addressing these disparities through targeted interventions, improved resource distribution, and investment in training will be crucial for improving the overall service delivery in the country.

In addition, the PCA provided valuable insights into the multifaceted factors influencing health service readiness in Ethiopia. By identifying the key determinants—availability of supplies, trained personnel, infrastructure, and adherence to guidelines—the PCA highlights the critical areas where interventions are needed to improve the quality of health services. Addressing these issues will require both targeted policy interventions at the regional level and system-wide reforms to ensure equitable access to quality maternal, child, and family health services across Ethiopia.

Future studies could explore the impact of resource allocation strategies and management reforms on service readiness to identify best practices that can be scaled up across Ethiopia and similar settings in sub-Saharan Africa.

## Data Availability

All data used for this study can be accessed on the DHS Program website at https://dhsprogram.com/data/dataset/Ethiopia_SPA_2021.cfm?flag=0

## Acknowledgments

The authors wish to thank Sara Riese for her valuable input on this analysis. We also thank the Ethiopian Public Health Institute (EPHI) for graciously allowing us access to the Ethiopia Service Provision Assessment datasets.

## >Recommended citation

Church, Rachael L., Girma F., Mussema, Y., Abelti, G., A., Shewarega, 2025. *Assessing Factors Affecting Readiness for Reproductive, Maternal, Newborn, Child, and Adolescent Health Services Ethiopia SPA Survey 2021-22*.

## APPENDIX

**Appendix Table 1.**
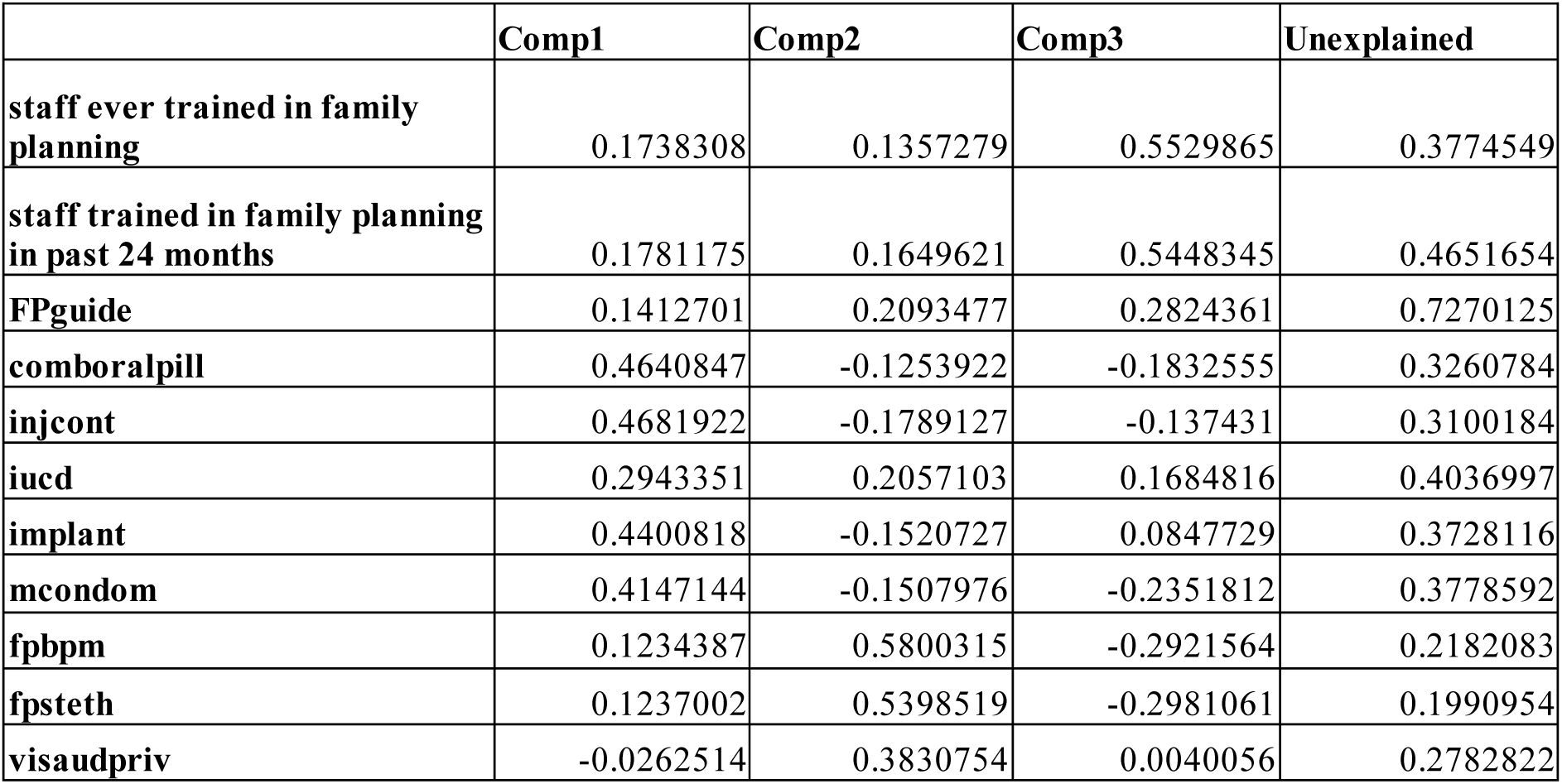
: Family Planning service readiness principal components factor loadings (eigenvectors)

**Appendix Table 2:**
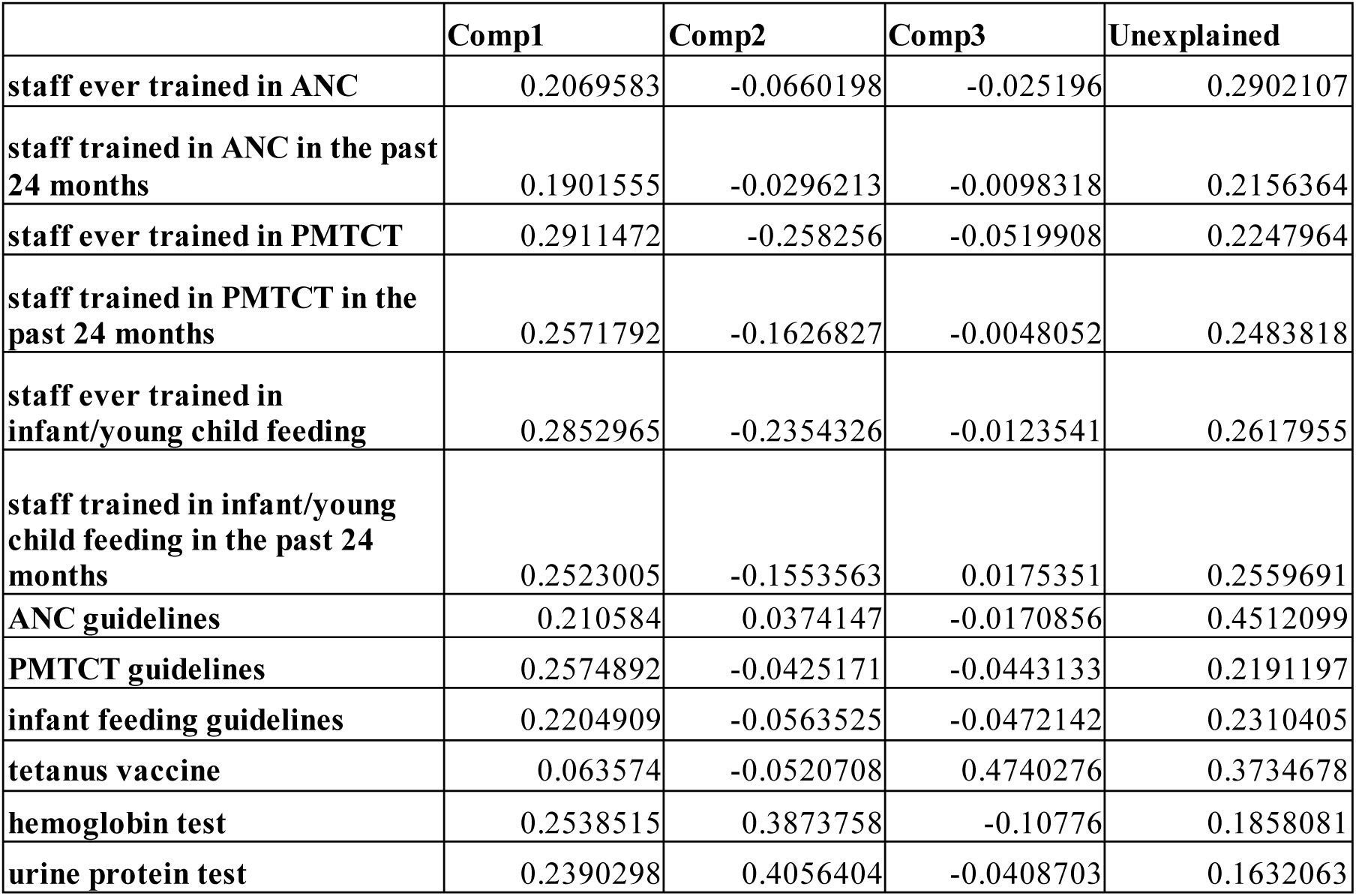
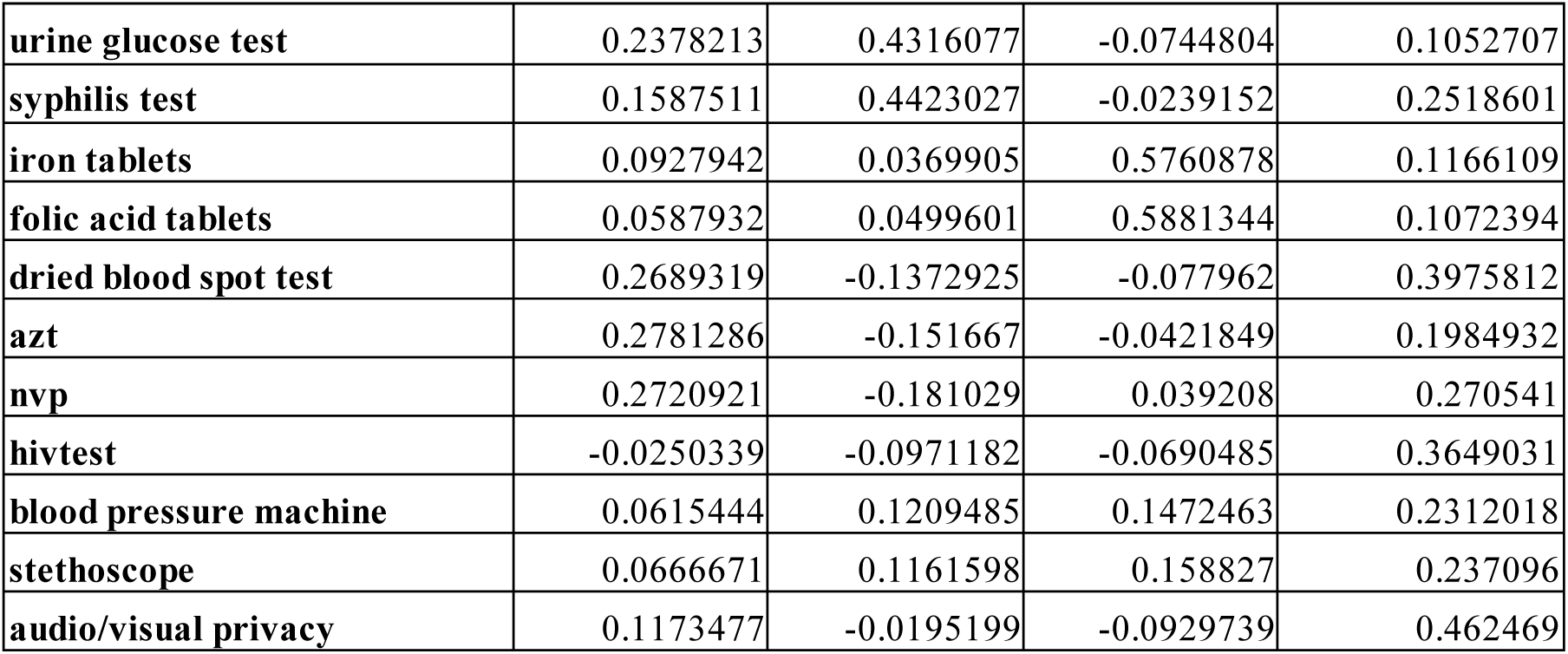
ANC/PMTCT service readiness principal components factor loadings (eigenvectors)

**Appendix Table 3:** BEmONC service readiness principal components factor loadings (eigenvectors)

|  | Comp1 | Comp2 | Comp3 | Unexplained |
| --- | --- | --- | --- | --- |
| staff ever trained in IMPAC | 0.0390563 | 0.6258325 | 0.2359194 | 0.2913871 |
| staff trained in IMPAC in the past 24 months | 0.0064513 | 0.6402536 | 0.0778763 | 0.3140321 |
| IMPAC guidelines | 0.2826558 | 0.1799534 | -0.0271057 | 0.5391173 |
| oxytocin | 0.2858523 | -0.1523992 | 0.39893 | 0.3577592 |
| diazepam | 0.2913145 | -0.0917296 | 0.0205319 | 0.3715266 |
| tetracycline | 0.3139369 | -0.0541559 | 0.2278521 | 0.4970415 |
| ceftriaxone | 0.2854178 | 0.0386788 | 0.0105188 | 0.5733694 |
| magnesium sulphate | 0.2844675 | -0.0469577 | 0.3640323 | 0.460266 |
| IV solution | 0.1965468 | -0.2116205 | 0.0425432 | 0.6096667 |
| exam light | 0.1330175 | 0.1268955 | 0.0501824 | 0.8578206 |
| suction apparatus | 0.214396 | 0.1504037 | -0.4491753 | 0.4836391 |
| manual vacuum extractor | 0.3213086 | 0.1089297 | -0.2063139 | 0.4640337 |
| vacuum aspirator kit | 0.2873402 | 0.0208956 | -0.3368789 | 0.3794123 |
| neonatal bag & mask | 0.2309201 | -0.1573729 | 0.14825 | 0.4299778 |
| fetal stethoscope | 0.1236414 | -0.0775306 | -0.2025921 | 0.3642784 |
| newborn corner equipment | 0.2791822 | 0.0333366 | 0.1045481 | 0.4565662 |
| neonatal resuscitation kit | 0.237432 | -0.0073661 | -0.3966582 | 0.4511363 |

**Appendix Table 4:** CEmOC service readiness principal components factor loadings (eigenvectors)

|  | Comp1 | Comp2 | Comp3 | Unexplained |
| --- | --- | --- | --- | --- |
| staff ever trained in CEmOC | 0.0805438 | 0.6310898 | 0.0364843 | 0.2864023 |
| staff trained in CEmOC in the past 24 months | -0.0223194 | 0.6500427 | 0.1295095 | 0.3195206 |
| CEmOC guidelines | -0.1230804 | 0.0448886 | 0.502369 | 0.3100228 |
| incubator | -0.1482113 | 0.1521883 | 0.2853334 | 0.3758832 |
| other heat source | 0.083509 | -0.0237673 | 0.451087 | 0.2272007 |
| anesthesia machine | 0.0905624 | 0.3506281 | -0.2161791 | 0.3762835 |
| ventilator vaporizers | -0.0307937 | 0.1298287 | -0.3051139 | 0.3342291 |
| anesthesia trolley | 0.3326705 | -0.0870642 | 0.3576658 | 0.2960348 |
| blood transfusions offered | 0.5533198 | -0.0027657 | -0.063603 | 0.1035162 |
| acceptable blood source | 0.5651017 | -0.0411624 | -0.0504799 | 0.0886486 |
| anesthetist | 0.2620612 | 0.0655964 | -0.3062726 | 0.5345017 |
| surgeon | 0.3673243 | 0.0069497 | 0.2765463 | 0.3057371 |

**Appendix Table 5:** Child vaccination service readiness principal components factor loadings (eigenvectors)

|  | Comp1 | Comp2 | Comp3 | Unexplained |
| --- | --- | --- | --- | --- |
| staff ever trained in child vaccination | 0.0229144 | 0.3793241 | -0.5830944 | 0.3053728 |
| staff trained in child vaccination in the past 24 months | 0.0414543 | 0.3627198 | -0.5827763 | 0.3277803 |
| child vaccination guidelines | -0.0387827 | 0.2090854 | 0.3313795 | 0.3368541 |
| pentavalent | 0.4530353 | 0.0025983 | 0.0406284 | 0.044355 |
| oral polio vaccine | 0.4424903 | 0.0095342 | -0.0291767 | 0.088068 |
| measles vaccine | 0.45238 | -0.0026833 | 0.0420813 | 0.0452579 |
| BCG vaccine | 0.4427093 | 0.0114644 | 0.0253128 | 0.0857569 |
| refrigerator | 0.4295516 | -0.0216094 | 0.0332246 | 0.133617 |
| vaccine carrier | 0.0926293 | 0.1457435 | 0.158002 | 0.344805 |
| sharps box | -0.0361253 | 0.6090055 | 0.2781446 | 0.3306433 |
| syringes and needles | -0.013219 | 0.5366551 | 0.3197091 | 0.4003042 |

**Appendix Table 6:**
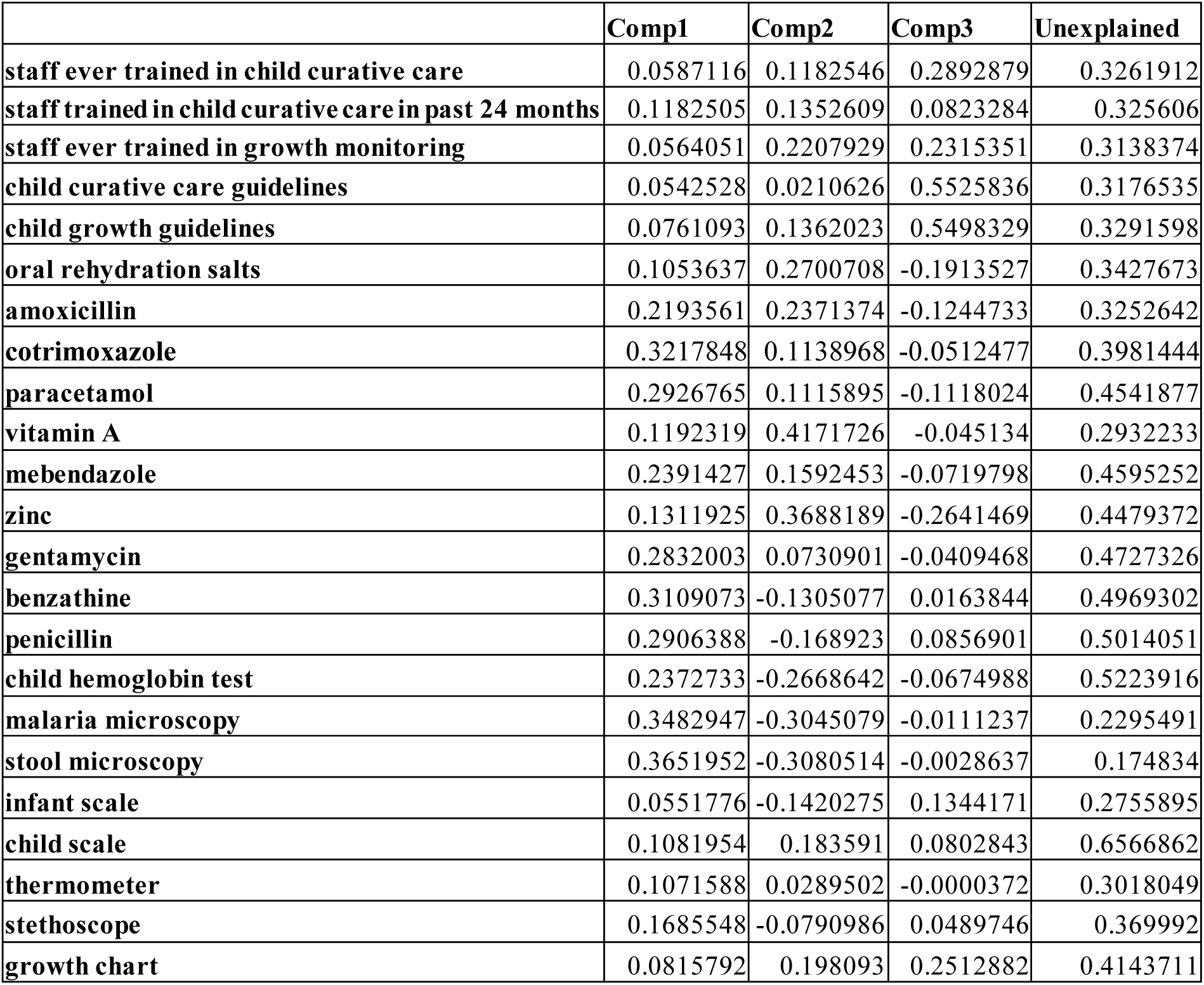
Child curative care and growth monitoring service readiness principal components factor loadings (eigenvectors)

|  | Comp1 | Comp2 | Comp3 | Unexplained |
| --- | --- | --- | --- | --- |
| staff ever trained in child curative care | 0.0587116 | 0.1182546 | 0.2892879 | 0.3261912 |
| staff trained in child curative care in past 24 months | 0.1182505 | 0.1352609 | 0.0823284 | 0.325606 |
| staff ever trained in growth monitoring | 0.0564051 | 0.2207929 | 0.2315351 | 0.3138374 |
| child curative care guidelines | 0.0542528 | 0.0210626 | 0.5525836 | 0.3176535 |
| child growth guidelines | 0.0761093 | 0.1362023 | 0.5498329 | 0.3291598 |
| oral rehydration salts | 0.1053637 | 0.2700708 | -0.1913527 | 0.3427673 |
| amoxicillin | 0.2193561 | 0.2371374 | -0.1244733 | 0.3252642 |
| cotrimoxazole | 0.3217848 | 0.1138968 | -0.0512477 | 0.3981444 |
| paracetamol | 0.2926765 | 0.1115895 | -0.1118024 | 0.4541877 |
| vitamin A | 0.1192319 | 0.4171726 | -0.045134 | 0.2932233 |
| mebendazole | 0.2391427 | 0.1592453 | -0.0719798 | 0.4595252 |
| zinc | 0.1311925 | 0.3688189 | -0.2641469 | 0.4479372 |
| gentamycin | 0.2832003 | 0.0730901 | -0.0409468 | 0.4727326 |
| benzathine | 0.3109073 | -0.1305077 | 0.0163844 | 0.4969302 |
| penicillin | 0.2906388 | -0.168923 | 0.0856901 | 0.5014051 |
| child hemoglobin test | 0.2372733 | -0.2668642 | -0.0674988 | 0.5223916 |
| malaria microscopy | 0.3482947 | -0.3045079 | -0.0111237 | 0.2295491 |
| stool microscopy | 0.3651952 | -0.3080514 | -0.0028637 | 0.174834 |
| infant scale | 0.0551776 | -0.1420275 | 0.1344171 | 0.2755895 |
| child scale | 0.1081954 | 0.183591 | 0.0802843 | 0.6566862 |
| thermometer | 0.1071588 | 0.0289502 | -0.0000372 | 0.3018049 |
| stethoscope | 0.1685548 | -0.0790986 | 0.0489746 | 0.369992 |
| growth chart | 0.0815792 | 0.198093 | 0.2512882 | 0.4143711 |

**Appendix Table 7:**
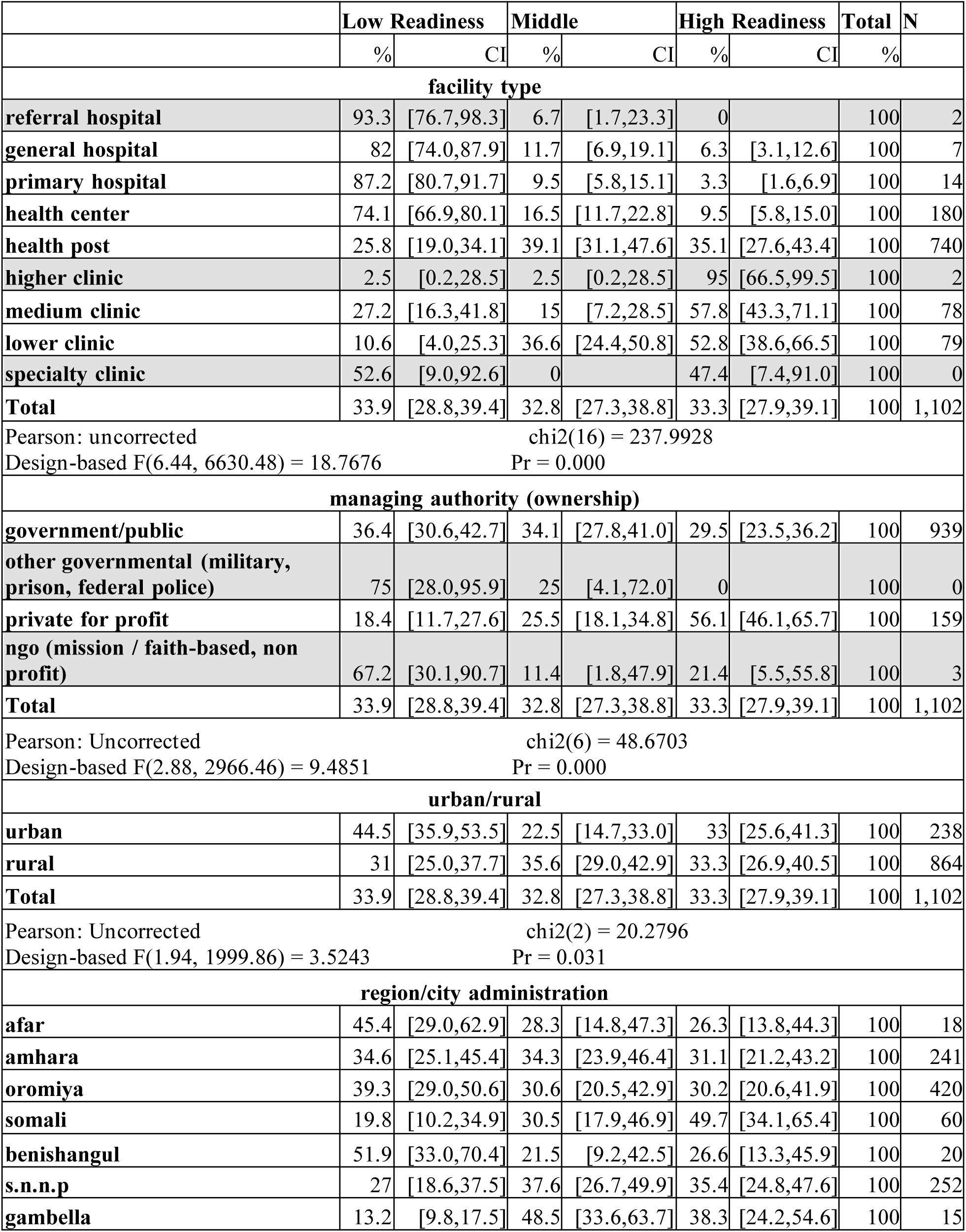
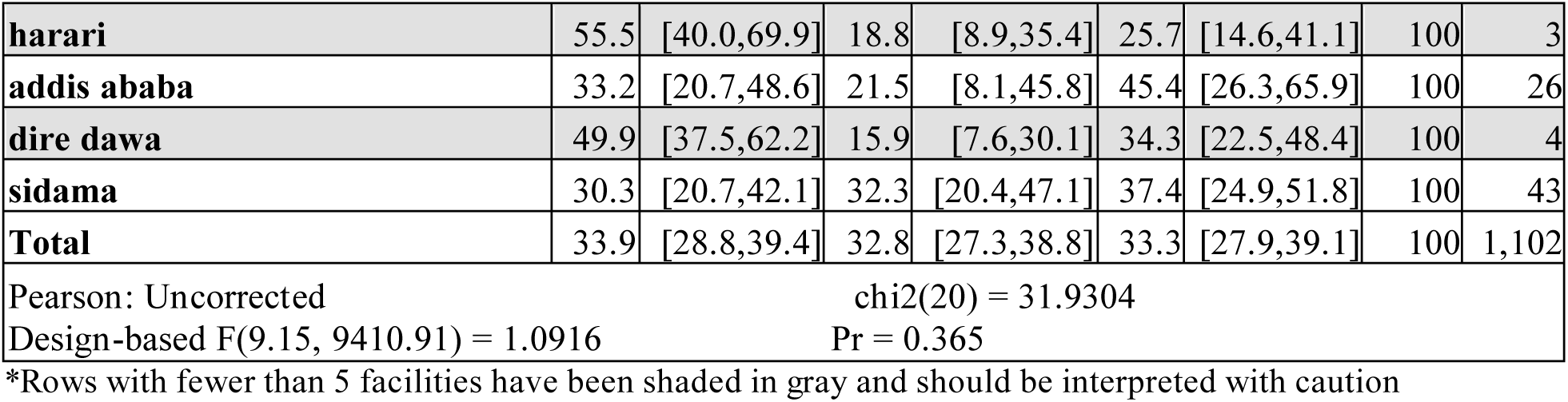
Family Planning Service Readiness by all facility characteristics in Ethiopia (2021-22)

**Appendix Table 8:**
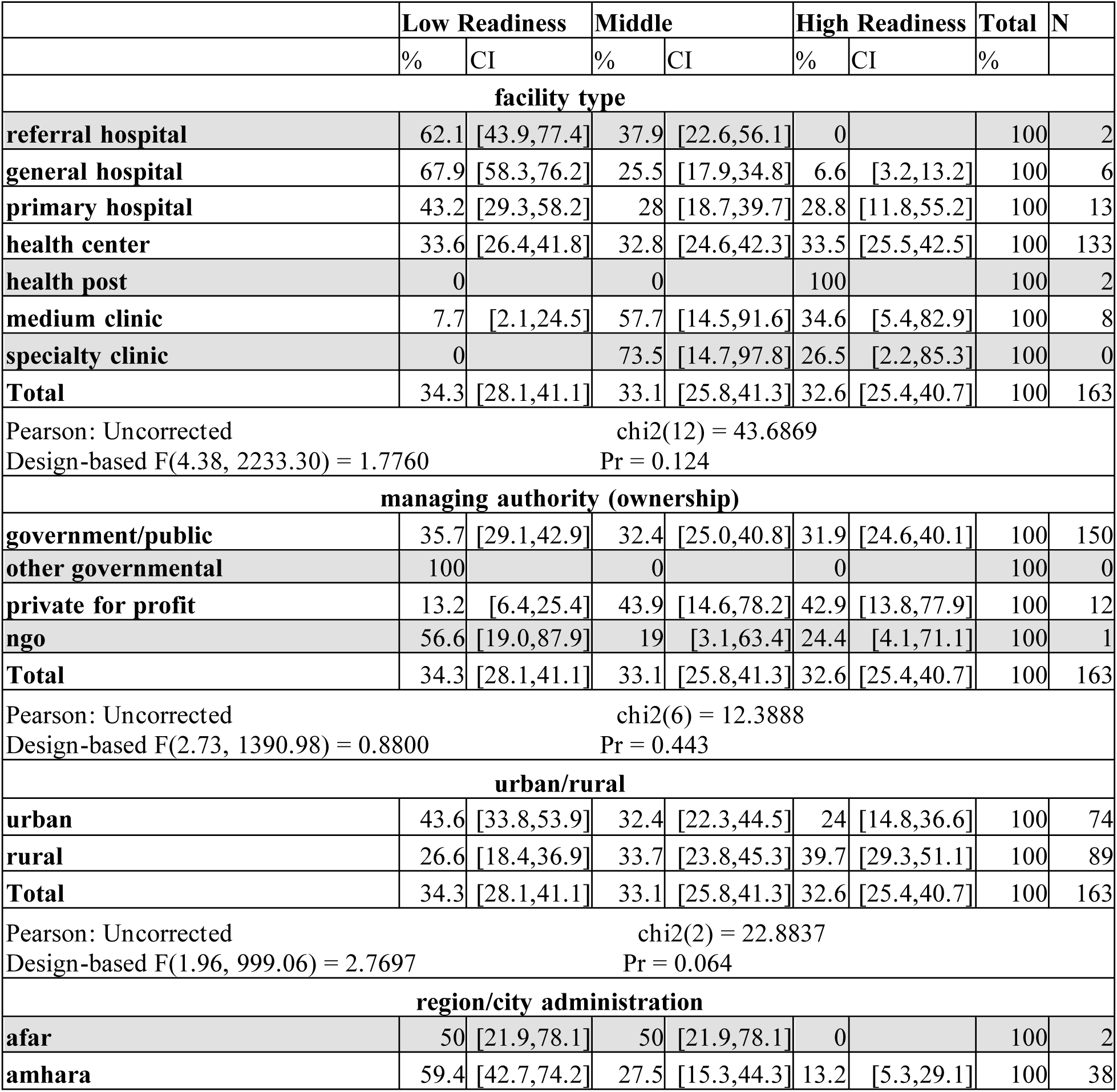
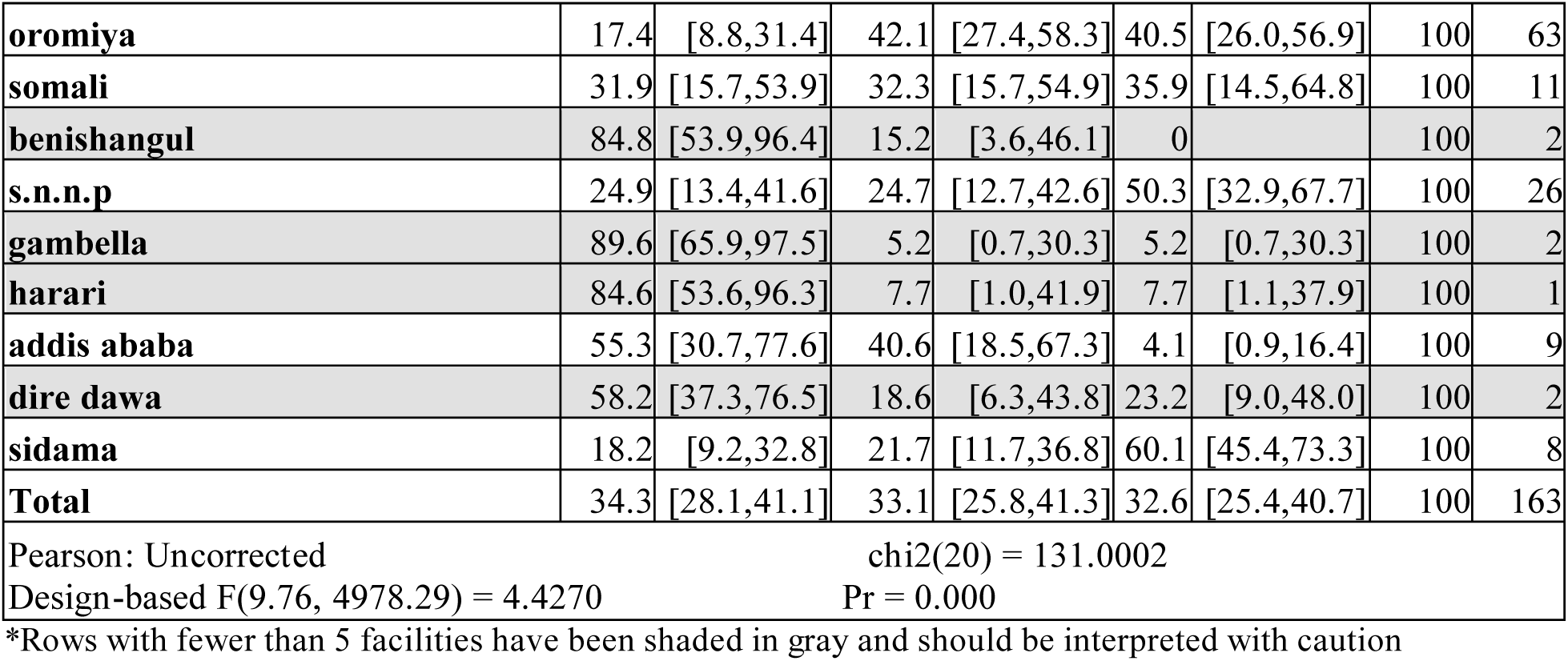
ANC and PMTCT Service Readiness by all facility characteristics in Ethiopia (2021-22)

**Appendix Table 9:**
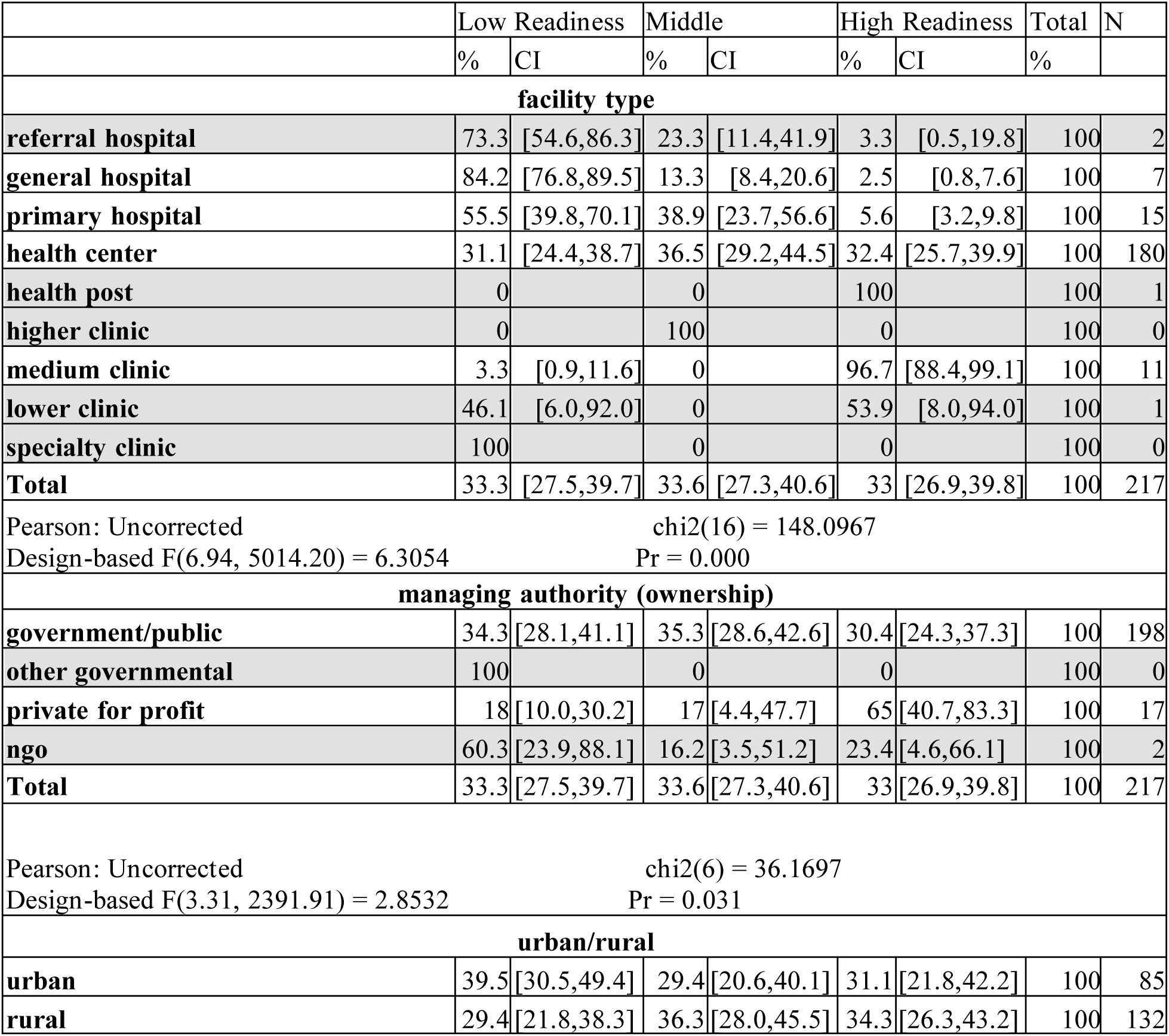
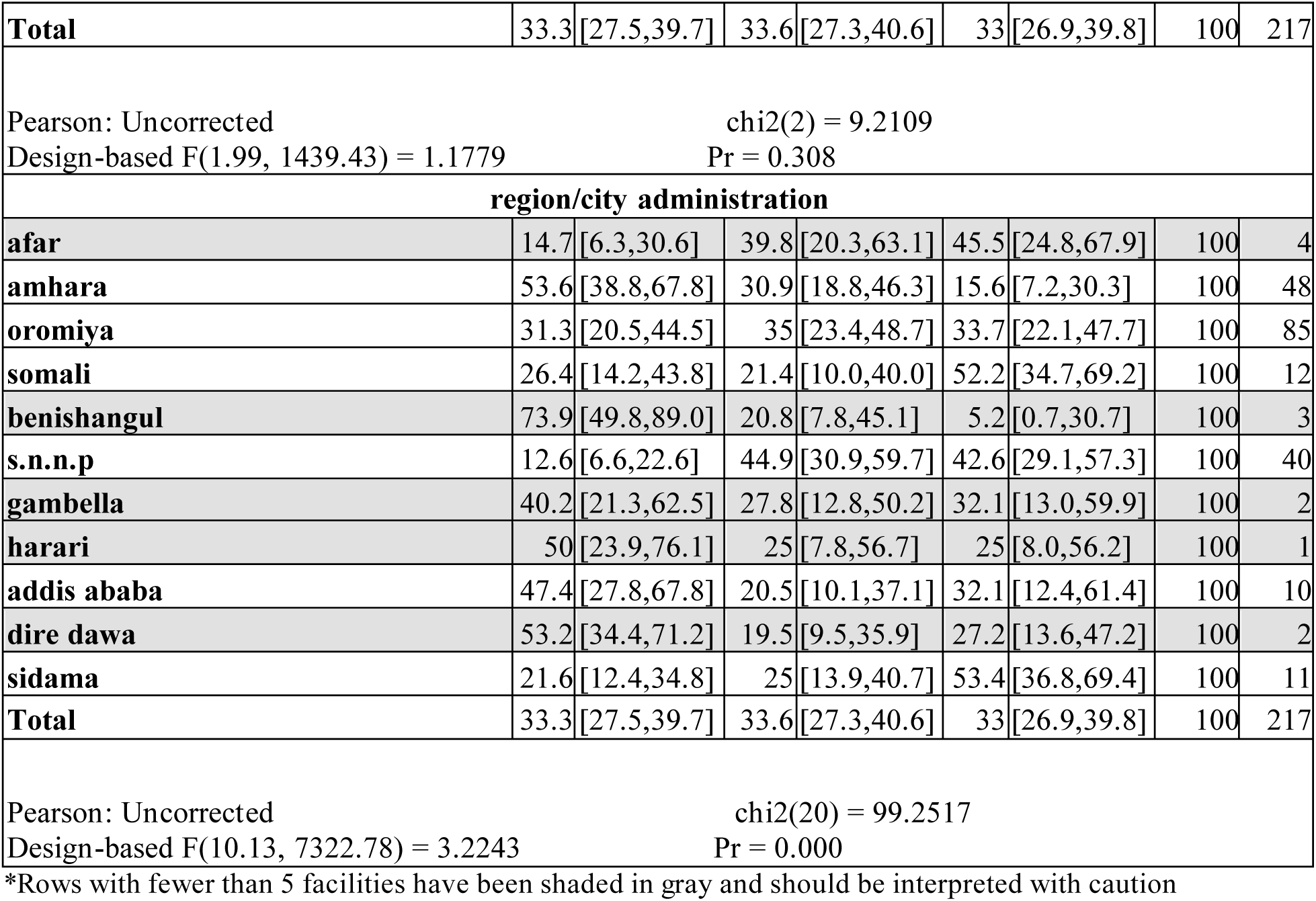
BEmONC Service Readiness by all facility characteristics in Ethiopia (2021-22)

**Appendix Table 10:**
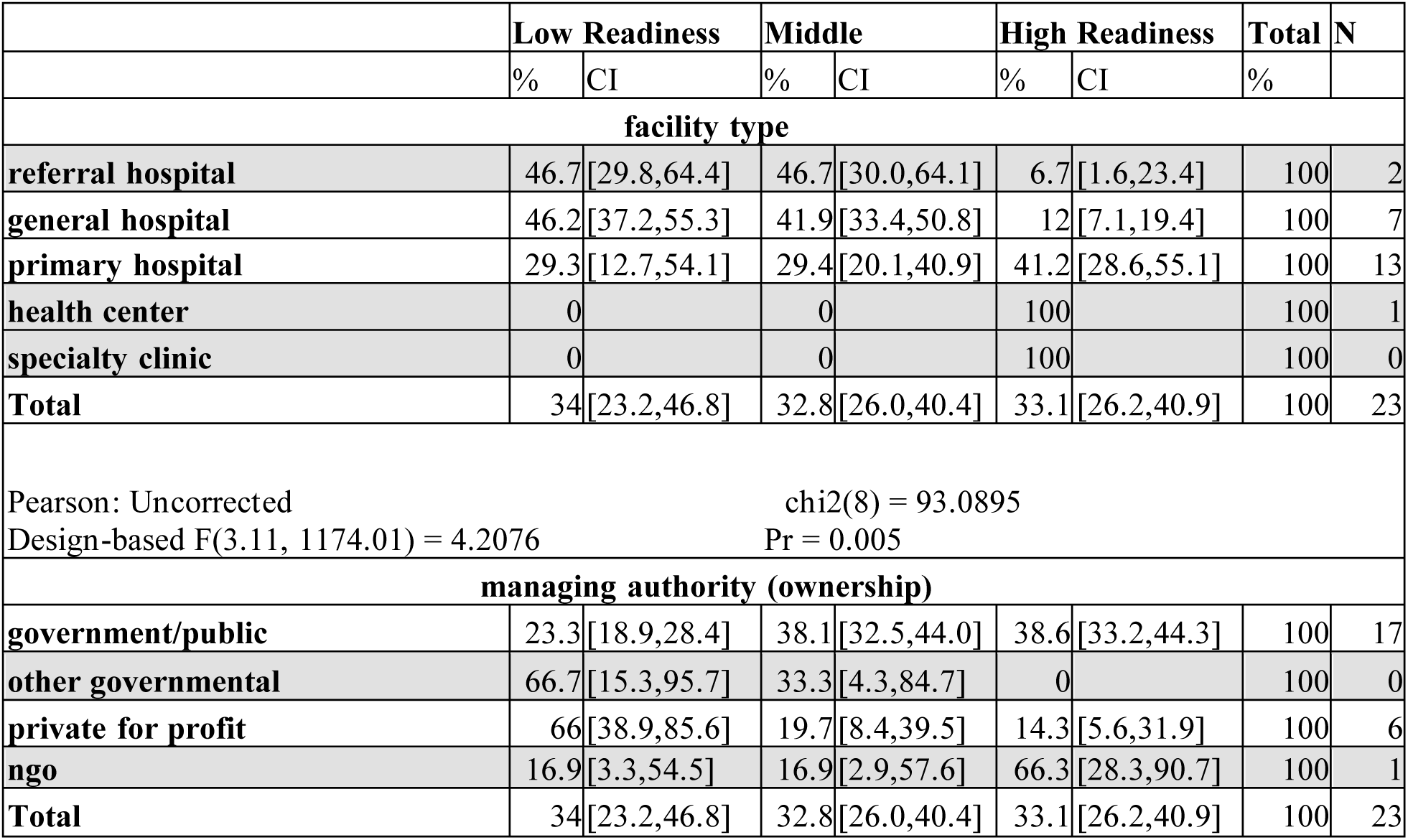
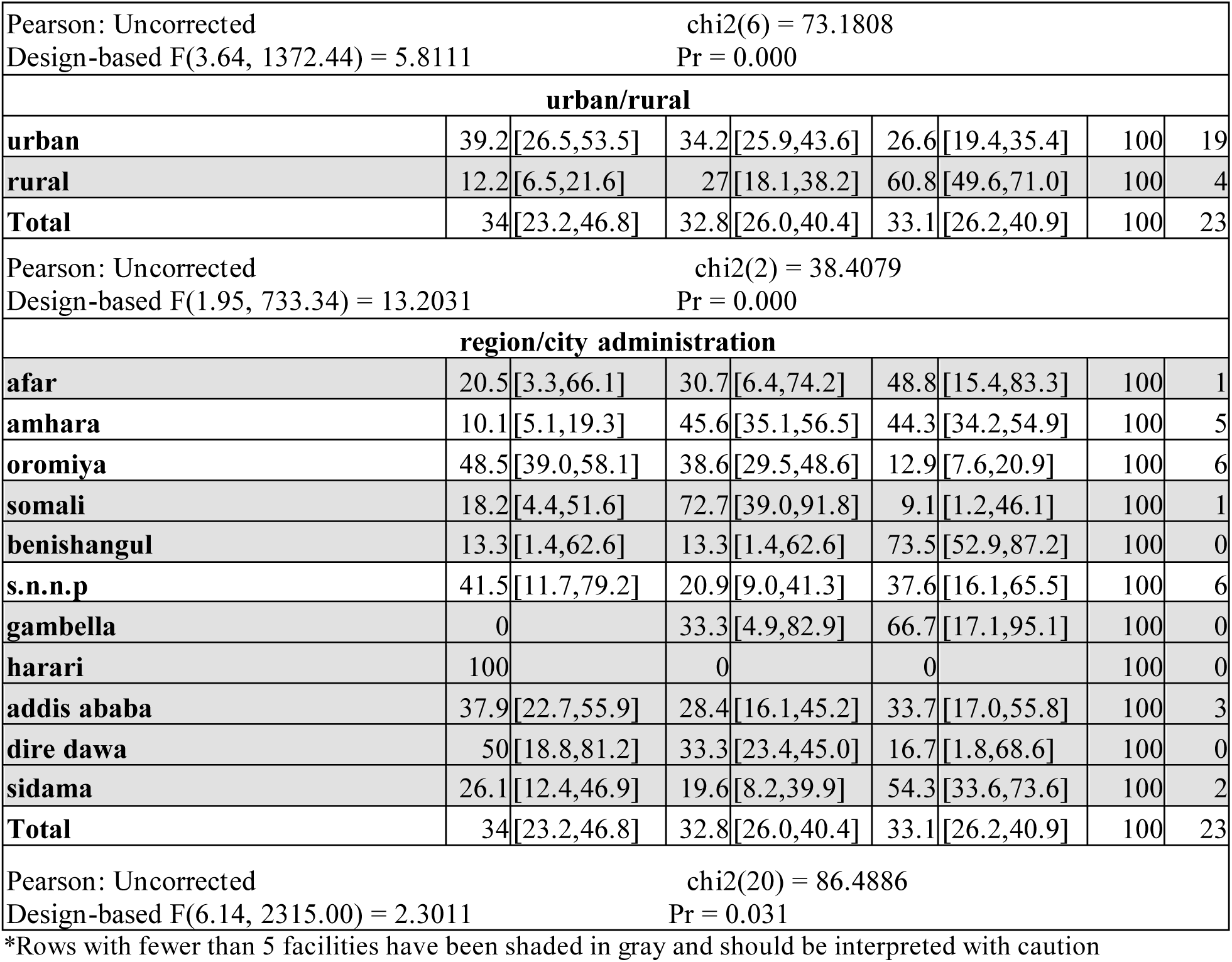
CEmOC Service Readiness by all facility characteristics in Ethiopia (2021-22)

**Appendix Table 11:**
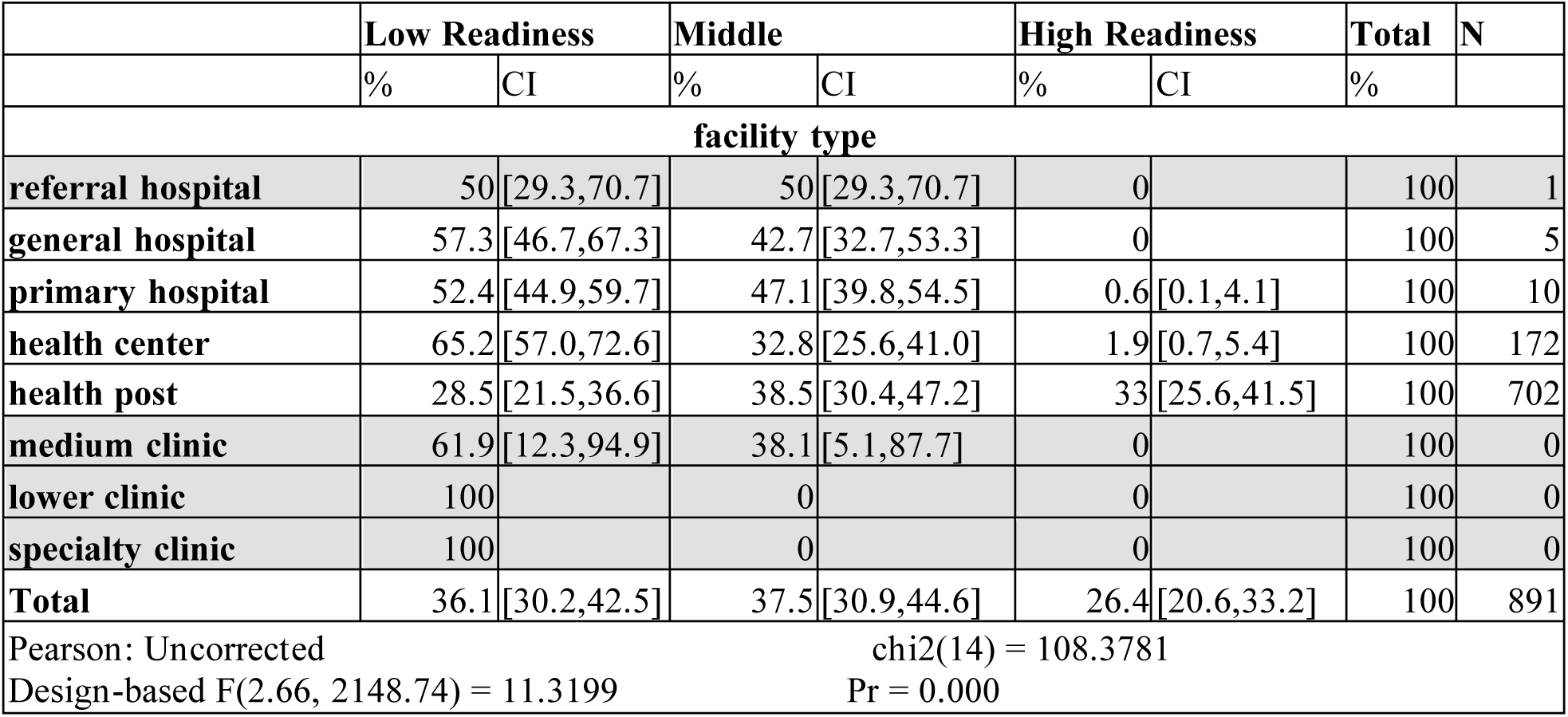
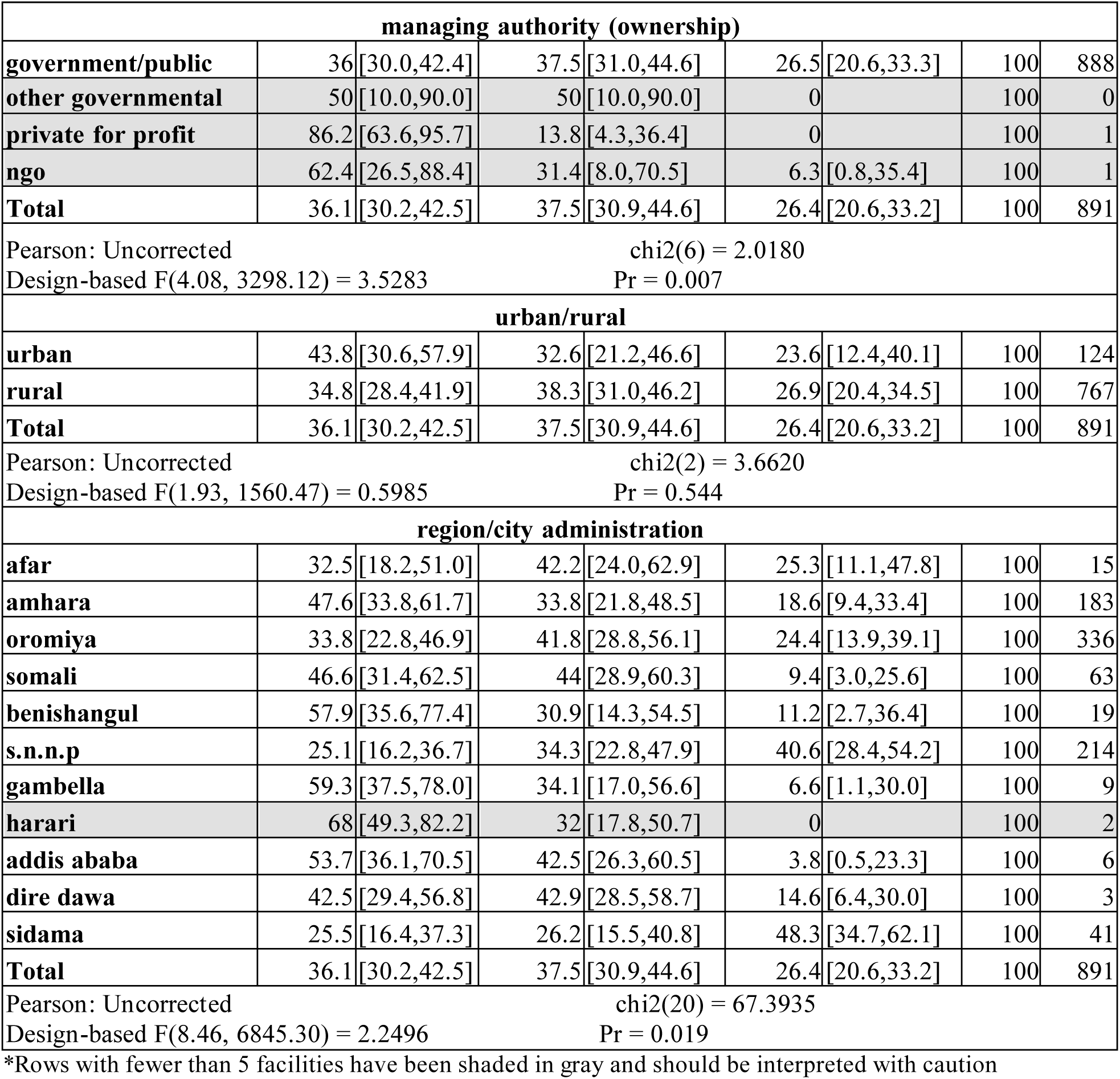
Child Vaccination Service Readiness by all facility characteristics in Ethiopia (2021-22)

**Appendix Table 12:**
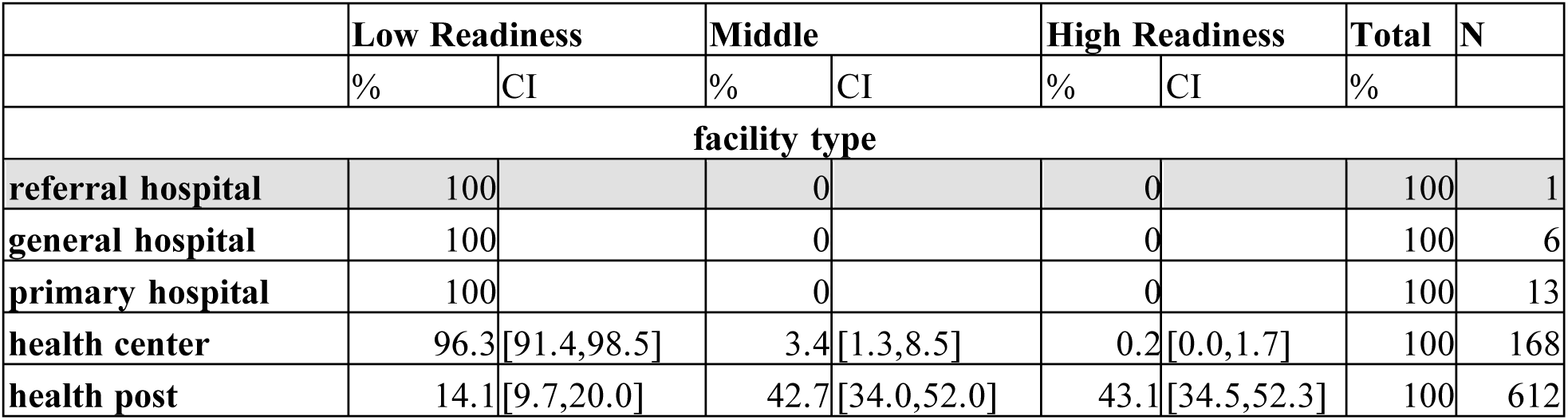
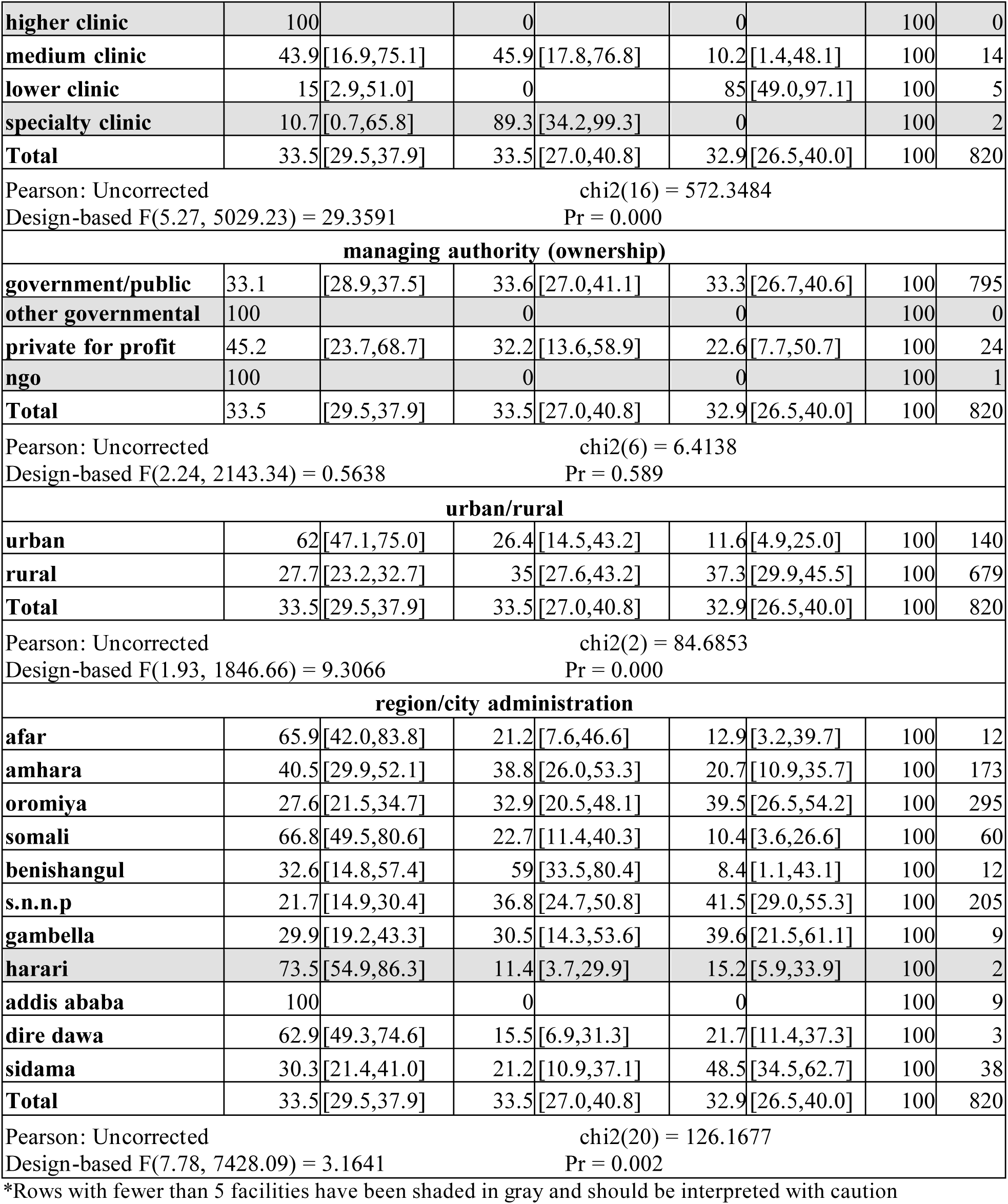
Child Curative Care and Growth Monitoring Service Readiness by all facility characteristics in Ethiopia (2021-22)

## Notes

### Competing Interest Statement

The authors have declared no competing interest.

### Funding Statement

All authors of this manuscript were formerly associated with the Demographic and Health Surveys Program through their respective institutions (Avenir Health, ICF, and USAID Ethiopia) and lost their jobs as a result of the shut down of USAID. We began the work on this project at our prior employment under the DHS Program and USAID, but at the termination of our roles decided to complete the manuscript on our own time, independently of our former affiliations and without any financial support. All data included are publicly available and therefore support from any institution was not needed to continue with publishing the manuscript.

### Author Declarations

IRB bodies were not engaged as this analysis did not include human subjects research.

## REFERENCES

1. *Ethiopia Service Provision Assessment 2021–22 Final Report*. Ethiopian Public Health Institute (EPHI), Ministry of Health, and ICF, 2023.

2. *Trends in maternal mortality 2000 to 2023: estimates by WHO, UNICEF, UNFPA, World Bank Group, and UNDESA/Population Division.* World Health Organization, 2025.

3. *Levels and trends in child mortality United Nations Inet-Agency Group for Child Mortality Estimation (UN IGME), Report* 2024. United Nations Inter-agency Group for Child Mortality Estimation, 2025.

4. Ensure healthy lives and promote well-being for all at all ages. United Nations Department of Economic and Social Affairs, Sustainable Development, 2025.

5. Ethiopia Demographic and Health Survey 2005. Central Statistical Agency [Ethiopia] and ORC Macro, 2006.

6. Ethiopia Demographic and Health Survey 2024–25: Key Indicators. Ethiopian Statistical Service (ESS) and ICF, 2025.

7. *Sustainable Development Goals.* United Nations. https://sdgs.un.org/goals [Accessed February 11, 2026]

8. Mekonnen, Y., et al. “Improved antenatal care services in rural Ethiopia’s public health centers through the Enhancing Nutrition and Antenatal Infection Treatment (ENAT) intervention.” BMC Health Serv Res 25, 662. 2025.

9. Abebe, S., et al. “Decentralizing and task sharing within the primary health system improved access and quality of ANC services in Amhara and Oromia regions: pre-post health facility data.” BMC Prim. Care 25, 411. 2024.

10. Ayehu, T., et al. “Facility readiness and experience of women and health care providers in receiving and delivering obstetric care in comprehensive health posts in Ethiopia: a mixed method study.” BMC Health Serv Res 25, 303. 2025.

11. Trends in maternal mortality estimates 2000 to 2023: estimates by WHO, UNICEF, UNFPA, World Bank Group and UNDESA/Population Division. Geneva: World Health Organization; 2025. https://www.unfpa.org/sites/default/files/pub-pdf/9789240108462-eng.pdf

12. *Ethiopia Service Delivery Indicators Health Survey 2018-*2019. World Bank, 2019.

13. *Health sector transformation plan II.* Ethiopia Ministry of Health, 2021.

14. *Monitoring the building blocks of health systems: a handbook of indicators and their measurement strategies*. World Health Organization. 2010.

15. Mallick, L., Wenjuan W., and Gheda T. A Comparison of Summary Measures of Quality of Service and Quality of Care for Family Planning in Haiti, Malawi, and Tanzania. DHS Methodological Report No. 20. ICF, 2017.

16. Kaiser, H. F. “An index of factorial simplicity.” Psychometrika, 39(*1*), 31–36, 1974. 10.1007/BF02291575

17. *Ethiopian Essential Health Services Package.* Ethiopia Ministry of Health. 2019.

18. Gakidou, E., et al. "The impact of private health insurance on health care utilization and quality of care in Kenya." The Lancet Global Health, 2016.

19. Benson, L., et. al. “Society of Family Planning interim clinical recommendations: Contraceptive provision when healthcare access is restricted due to pandemic response.” Society of Family Planning, 2016. 10.46621/UYGR2287

20. Ameh, C., et al. "Barriers to the provision of child immunization services in Nigeria.per" The Lancet, 2017.

21. Chola, L., et al. "Service readiness and equity in maternal and child health services in South Africa." BMC Public Health, 2020.

22. *Trends in maternal mortality 2000 to 2020.* World Health Organization, 2021.

23. Nabagereka, M., et al. "Urban health systems and service delivery in Uganda." Uganda Journal of Health Systems, 2018.

24. Juma, E., et al. "Health system challenges in providing basic emergency obstetric and neonatal care in Tanzania." Tanzania Journal of Health Research, 2017.

25. Mugisha, F., et al. "Private versus public hospitals: The quality of maternal health services in Tanzania." BMC Health Services Research, 2020.

26. Kondwani, S. A., et al. "The challenges faced by health centers in providing curative care services in South Africa." South African Medical Journal, 2021.

